# Evaluating the effects of SARS-CoV-2 Spike mutation D614G on transmissibility and pathogenicity

**DOI:** 10.1101/2020.07.31.20166082

**Authors:** Erik Volz, Verity Hill, John T. McCrone, Anna Price, David Jorgensen, Áine O’Toole, Joel Southgate, Robert Johnson, Ben Jackson, Fabricia F. Nascimento, Sara M. Rey, Samuel M. Nicholls, Rachel M. Colquhoun, Ana da Silva Filipe, James Shepherd, David J. Pascall, Rajiv Shah, Natasha Jesudason, Kathy Li, Ruth Jarrett, Nicole Pacchiarini, Matthew Bull, Lily Geidelberg, Igor Siveroni, Ian Goodfellow, Nicholas J. Loman, Oliver G. Pybus, David L. Robertson, Emma C. Thomson, Andrew Rambaut, Thomas R. Connor, on behalf of the CoG-UK consortium

## Abstract

Global dispersal and increasing frequency of the SARS-CoV-2 Spike protein variant D614G are suggestive of a selective advantage but may also be due to a random founder effect. We investigate the hypothesis for positive selection of Spike D614G in the United Kingdom using more than 25,000 whole genome SARS-CoV-2 sequences. Despite the availability of a large data set, well represented by both Spike 614 variants, not all approaches showed a conclusive signal of positive selection. Population genetic analysis indicates that 614G increases in frequency relative to 614D in a manner consistent with a selective advantage. We do not find any indication that patients infected with the Spike 614G variant have higher COVID-19 mortality or clinical severity, but 614G is associated with higher viral load and younger age of patients. Significant differences in growth and size of 614G phylogenetic clusters indicate a need for continued study of this variant.

## Introduction

SARS-CoV-2, the coronavirus causing the global COVID-19 pandemic, is a rapidly-evolving RNA virus that continually accrues genomic mutations as it transmits. A major focus of current research into SARS-CoV-2 genetics is whether any of these mutations have the potential to significantly alter important viral properties, such as the mode or rate of transmission, or the ability to cause disease. Evolutionary theory predicts that most new viral mutations are deleterious and short-lived, whereas mutations that persist and grow in observed frequency may be either selectively neutral, or advantageous to viral fitness. Discriminating between these two scenarios and determining the selective benefit of new mutations is challenging, particularly for a newly-emergent virus such as SARS-CoV-2. For example, the observation that a new mutation is increasing in prevalence or geographic range is, by itself, insufficient to prove its selective advantage to the virus, because such increases can be generated by neutral epidemiological processes such as genetic bottlenecks following founder events and range expansions.

Considerable attention has focussed on the D614G mutation in SARS-CoV-2, a non-synonymous mutation resulting in a change from aspartic acid to glycine at position 614 of the virus’ S protein (D614G). The trimeric S protein, composed of subunits S1 and S2, is a large glycoprotein that mediates cell entry and has been studied extensively in other coronaviruses, including SARS-CoV (Belouzard et al., 2009; Li, 2015; Li et al., 2005) and MERS (Millet and Whittaker, 2014; Yang et al., 2014). SARS-CoV-2 S protein binds to angiotensin-converting enzyme 2 (ACE2) to gain cell entry, hence mutations in this gene have the potential to alter receptor binding affinity and infectivity, as well as viral immune evasion and immunogenicity(Watanabe et al., 2020).

The putative importance of the D614G mutation is based on three distinct sets of observations. First, experimental work using pseudotyped lentiviruses indicate that D614G increases infectivity *in vitro* (Korber et al., 2020; Yurkovetskiy et al.; Zhang et al., 2020). Second, structural analysis suggests that D614G alters the receptor binding conformation, such that ACE2 binding and fusion is more likely (Yurkovetskiy et al.). Third, analysis of the frequency of the 614D and 614G variants over time (based on submissions to global sequence databases) have suggested that locations which reported 614D viruses early in the pandemic were often later dominated by 614G viruses (Furuyama et al., 2020; Korber et al., 2020).

The D614G mutation is associated with the B.1 lineage of SARS-CoV-2 (Figure 1), which now dominates the global pandemic, based upon global SARS-CoV-2 genome sequences shared via GISAID (https://cov-lineaaes.ora/lineaaes/lineaae_B.1html. Retrospectively-sampled viruses suggest this mutation was present in Guangzhou, Sichuan and Shanghai Provinces, China in late January (Figure S1). In Europe, the 614G variant was first observed in genomes sampled on January 28th in a small outbreak in Bavaria, Germany, which was initiated by a visitor from Shanghai (Rothe et al., 2020) and subsequently controlled through public-health efforts. It is therefore likely that the D614G mutation occurred in China before being introduced on multiple occasions to European countries (Lai et al.) where it increased in frequency. This scenario is consistent with the rapid increase in February and March of European virus genomes that carry the 614G variant (Dearlove et al., 2020; Korber et al., 2020). In the UK, the first observation of a genome carrying the D614G mutation was in a sample collected on February 28th from a patient in Scotland who had recently travelled through Italy (Robertson, 2020).

**Figure 1.**
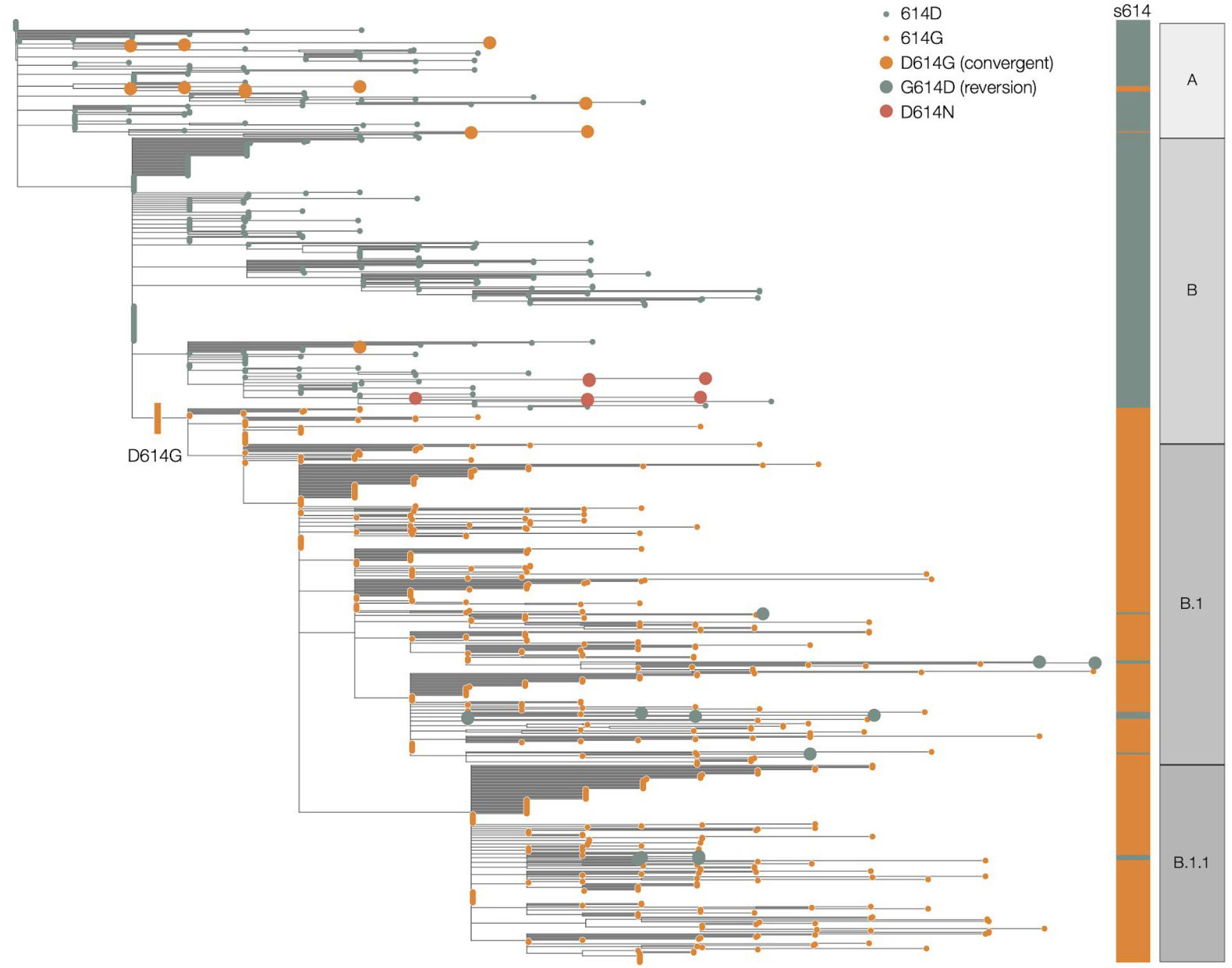
Maximum likelihood phylogeny estimated from a representative set of 900 SARS-CoV-2 genome sequences, showing global lineage assignments and the origins of the Spike protein D614G mutation which seeded many introductions in the United Kingdom. Putative reversions to 614D and independently arising D614G mutations are shown as large circles. The D614N genomes shown as red circles indicated two independent clusters in the UK.

There is currently no scientific consensus on the effect of the D614G mutation on SARS-CoV-2 infectivity and transmissibility, and some skepticism that it could produce a meaningful effect at the population level given that SARS-CoV-2 is already highly transmissible and rapidly spreading (van Dorp et al., 2020; Grubaugh et al., 2020). Although evaluating the effect of the D614G mutation on infectivity *in vitro* using pseudotype viruses is a critical first step, it is more challenging to determine whether any identified effects hold true for complete SARS-CoV-2 variants *in vitro* which requires an animal model that accurately reproduces meaningful aspects of virus infection and transmission. Even then, animal models may not accurately recapitulate the effect of variants on virus transmissibility within the human population. Therefore experimental evidence should be complemented with large-scale population studies that can detect meaningful changes in human-to-human transmission. The small size of the SARS-CoV-2 genomic datasets from many countries precludes robust analysis on a national scale. The substantially larger global SARS-CoV-2 sequence dataset is also problematic because of limited sequence metadata and variable sampling approaches among countries. To determine statistically if there is a meaningful difference in transmission between the 614D and 614G variants, we ideally need to observe repeated independent introductions of each variant into the same population and follow the trajectories of the outbreaks they cause.

In the UK, the rapid establishment of a national sequencing collaboration at the start of the epidemic in the UK, CoG-UK (The COVID-19 Genomics UK (COG-UK) consortium, 2020), has resulted in the generation of >40,000 SARS-CoV-2 sequences from the country in <6 months (approximately half of all genomes sequenced globally as of the 7th of July). CoG-UK has facilitated the usage of robust and systematic sampling, shared bioinformatic and laboratory approaches, and the collection of consistent core metadata, resulting in a large, high-resolution dataset capable of examining changes in virus biology in the UK. Crucially for this study, and in contrast to epidemics that followed the first European outbreaks, the UK epidemic is the result of repeated introduction of SARS-CoV-2 from numerous global locations, including a substantial number of phylogenetic sub-trees (clusters) carrying either 614D and 614G. Here we use the CoG-UK dataset to examine evidence for increased transmissibility of SARS-CoV-2 due to genetic changes in its Spike protein. We also investigate the influence of Spike 614D versus G on pathogenicity by matching sequence data with clinical outcome.

## Results

We identified 21,231 614G and 5,755 614D de-duplicated whole genome sequences sampled from different infections within the UK with known dates of sample collection between January 29 and June 16, 2020. We identified phylogenetic clusters of United Kingdom genomes using a maximum-parsimony reconstruction of the location of phylogenetic branches within the global SARS-CoV-2 phylogeny (see Methods). Each cluster stems from one or a small number of introductions of the virus into the UK. We identify 245 614G and 62 614D clusters containing UK virus genomes from 10 or more different patients, after removing samples with Spike 614 genotype which does not match the majority within their cluster (reversions or contaminations). Importantly, we identified more UK phylogenetic clusters carrying the 614G variant than the 614D variant, and on average the 614G clusters were first detected later (the mean detection date for 614G clusters was 16 days later than of 614D clusters; Figure 2). Whilst the frequency of sampling of 614G and 614D variants in the UK was close to parity in February and March, 614G became the dominant form in late March and this trend has continued (Figure 2C).

**Figure 2.**
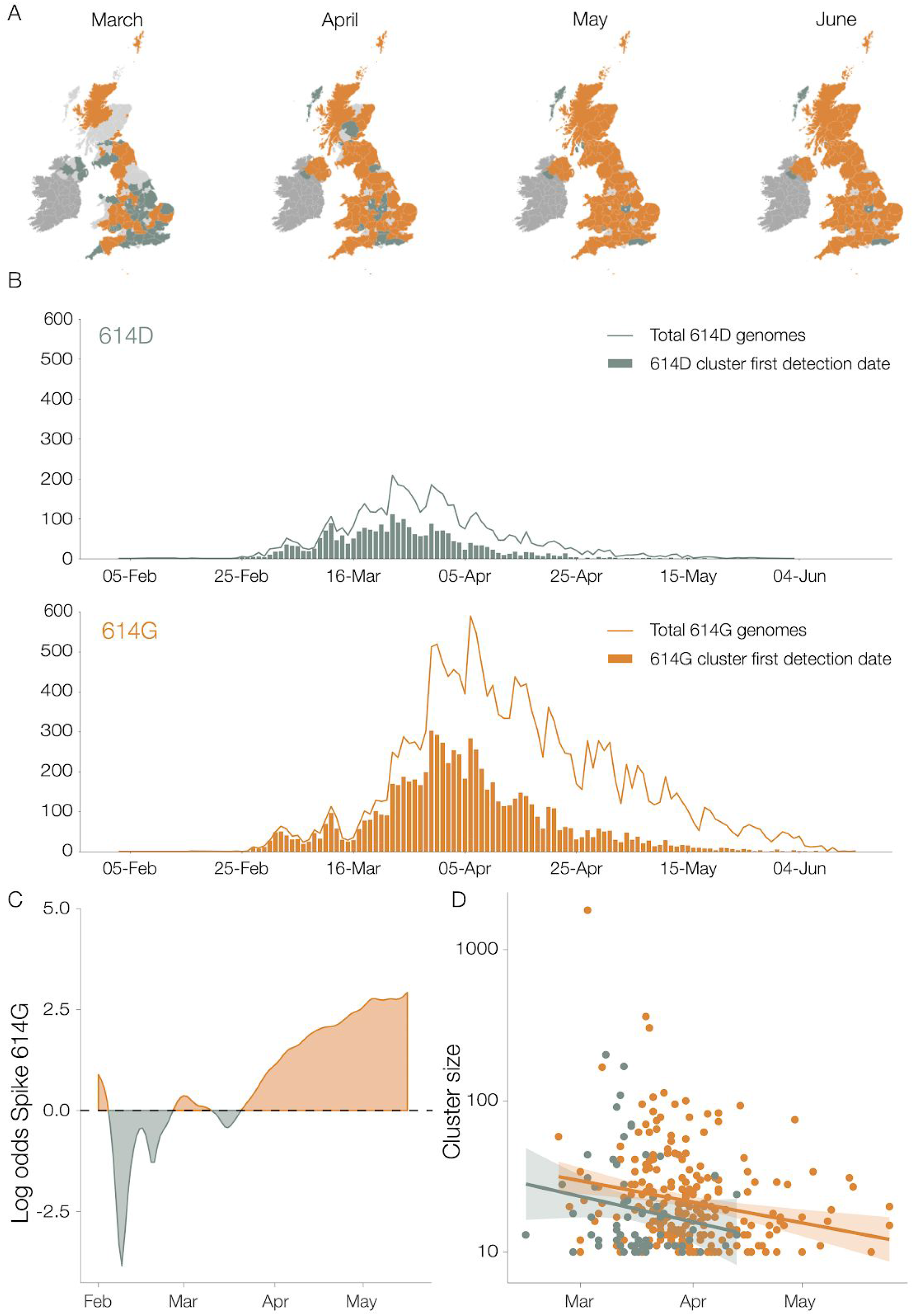
Geographic and temporal distribution of UK phylogenetic clusters, classified as 614D or 614G according to the residue they carry at S protein position 614D. A) Shaded regions show the predominant residue in each region on the 15th of each month for March, April, May and June 2020, with orange indicating that 614G was more frequently sampled and green indicating that 614D was more (or equally) frequent. Light grey indicates that no sequences had been sampled by that point in time. Dark grey indicates the Republic of Ireland. B) The date when each cluster was first detected in the United Kingdom, for variants 614D and 614G. Each cluster contains 2 or more sampled genomes. Solid lines show the total number of sequences collected by day of each 614 variant. C) The log odds of sampling a 614G variant over time. D) The size of cluster versus time of first sample collected within a cluster.

### Evaluating the hypothesis that 614G confers increased transmission fitness

UK phylogenetic clusters that were first detected early in the epidemic tend to be larger than those detected later (Figure 2D). Although most 614G clusters tended to be detected later, they are on average 59% larger than 614D clusters after adjusting for the time of cluster detection (*p*=0.008).

To evaluate if the increasing sampling frequency of 614G reflects a selective advantage, we fit a logistic growth model to observed sequence sampling dates, under the assumption that sequences are sampled in proportion to the relative frequency of the 614G variants in the population, which changes over time. Under this model, 614D-infected cases grow exponentially at a rate *r* and 614G-infected cases grow exponentially at rate 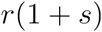, where *s* represents the estimated mutational selection coefficient.

In order to account for the rapid increase in SARS-CoV-2 introduction into the UK during March (Pybus et al., 2020) we adapted the logistic growth model to count only those sequences that belong to clusters first detected in January or February. We further limit the analysis to sequences sampled during a period of exponential growth up to the end of March shortly after a national lockdown was implemented in the UK. Origin times for clusters were estimated using molecular clock phylogenetic methods (c.f. Methods). We also only consider samples collected after the most recent common ancestor (TMRCA) of the individual TMRCA of all clusters and where there are at least 10 samples with *either* amino acids 614D or G. Under these conditions, all samples included in the analysis were collected during a period when the selected clusters were co-circulating within the UK.

Consequently, for this analysis, we retained 5 614D clusters (n=355 sequences) and 5 614G clusters (n=1,855 sequences) and estimated a selection coefficient for the 614G of 0.21 (95% CI: 0.06 – 0.56) (Table 1). The observed and fitted frequencies of 614G samples are shown in Figures 3A and S2. Information used to fit this model is drawn disproportionately from late March when more sequences are available.

**Table 1.**
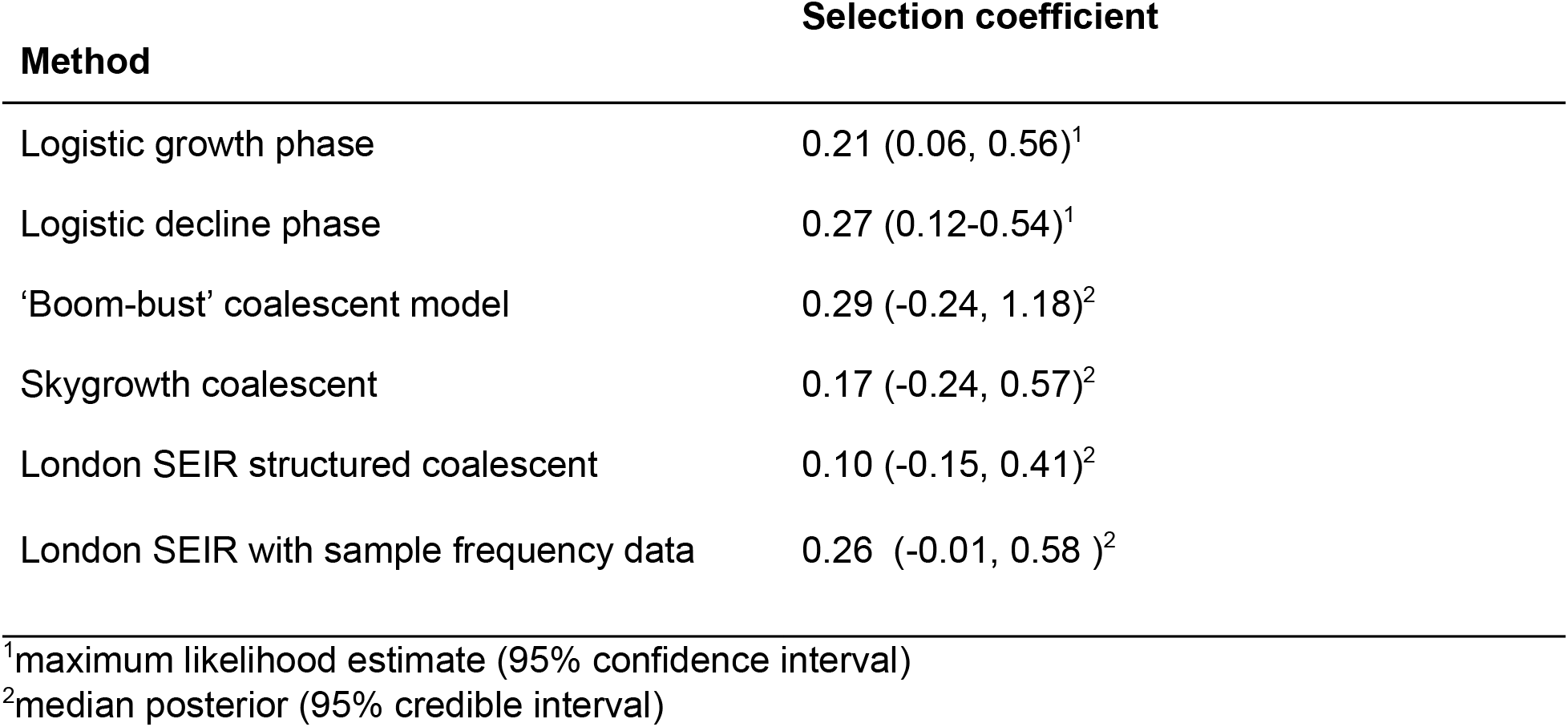
Estimates of the selection coefficient of the 614G variant using different data sets and models.

| Method | Selection coefficient |
| --- | --- |
| Logistic growth phase | 0.21 (0.06, 0.56)<sup>1</sup> |
| Logistic decline phase | 0.27 (0.12-0.54)<sup>1</sup> |
| 'Boom-bust' coalescent model | 0.29 (-0.24, 1.18)<sup>2</sup> |
| Skygrowth coalescent | 0.17 (-0.24, 0.57)<sup>2</sup> |
| London SEIR structured coalescent | 0.10 (-0.15, 0.41)<sup>2</sup> |
| London SEIR with sample frequency data | 0.26 (-0.01, 0.58)<sup>2</sup> |

<sup>1</sup>maximum likelihood estimate (95% confidence interval)
<sup>2</sup>median posterior (95% credible interval)

**Figure 3.**
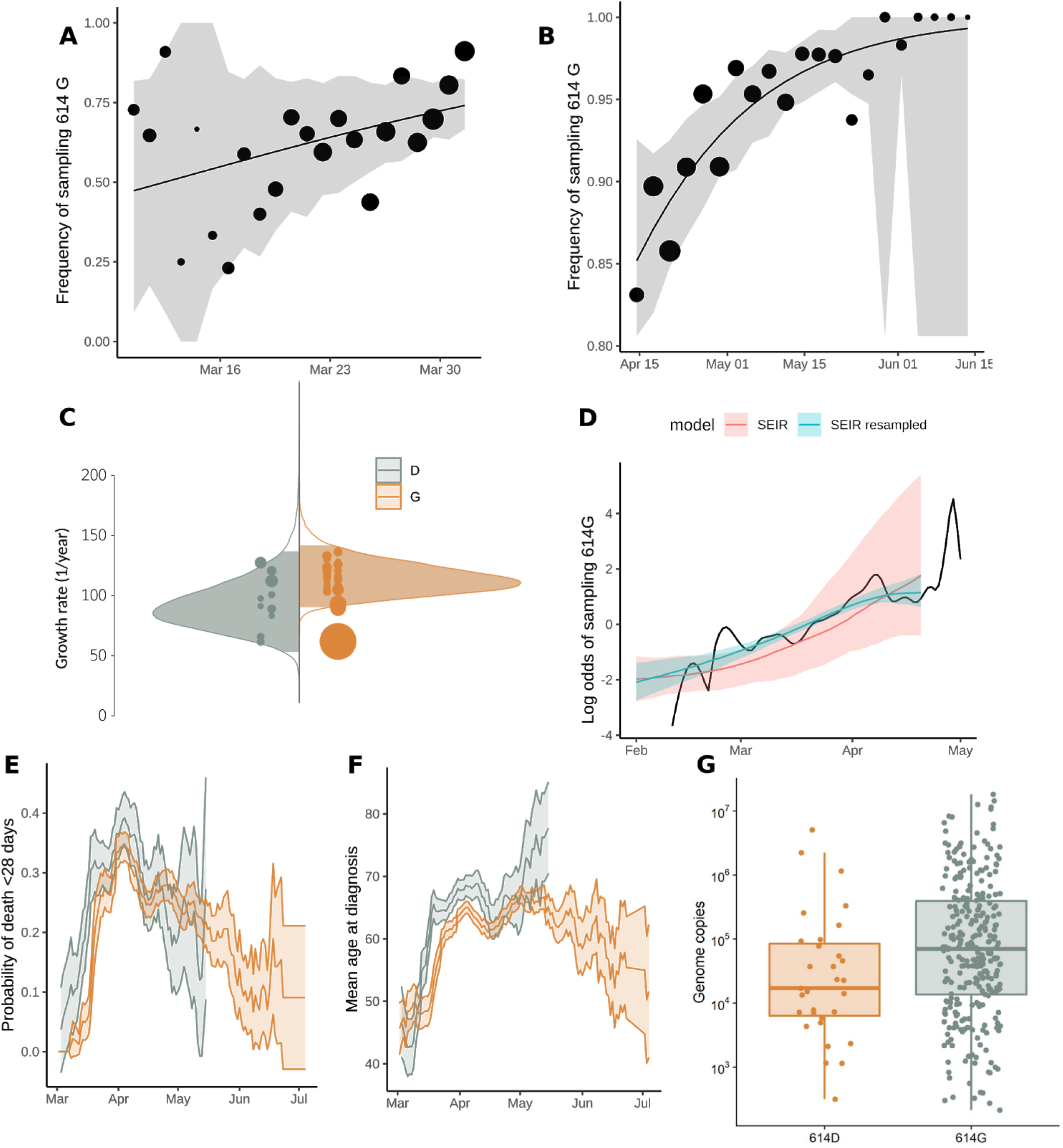
Relative frequency of Spike 614 D and G over time (A-B), phylodynamic growth rates (C-D), and comparison of clinical severity metrics (E-G). A) Frequency of sampling Spike 614G over time for clusters sampled during exponential growth phase. The size of points represents the number of samples collected on each day. The line and shaded region showed the MLE and confidence interval fit of the logistic growth model. B) As in (A) but including samples during a period after April 15 during a period of epidemic decline. C) Distribution of exponential growth rate for Spike 614G (brown) and 614D (grey) in units of 1/year. Solid areas span the 95% credible interval. Points indicate the rates estimated for specific clusters, and are sized by the number of sequences in that cluster. D) Log odds of sampling Spike 614G in London comparing empirical values (black line) and estimates based on the phylodynamic SEIR model (shaded regions). The green shaded region shows estimates making use of both genetic data and sample frequency data. E) The probability over time of fatal outcome within 28 days of diagnosis among UK patients with sequence data that can be matched to clinical records. Shaded regions show 95% confidence region of a 7-day moving average. Points with fewer than 20 observations are omitted. F) Moving average of age among samples included in (E). G) Viral load (RT-qPCR mean genome copies) estimated using SARS-CoV-2 RNA strands from 31 614D (614D) and 290 614G samples.

We separately fitted the logistic growth model to the period of epidemic decline after April 15. If we include all clusters first detected before March 31, then we have n=3,335 sequences (3,093 614G and 242 614D) sampled after April 15 and belonging to 37 phylogenetic clusters. This cross-section of data also exhibits a trend of an increasing frequency of 614G through time (Figures 3B and S3), with an estimated selection coefficient of 0.27 (95% CI:0.12-0.54).

An alternative source of information about the relative growth rates of the two variants comes from changing patterns of genetic diversity over time in each cluster. We applied phylodynamic methods (Pybus and Rambaut, 2009) to estimate effective population size and effective growth rates over time. First, we applied a parametric ‘boom-bust’ exponential growth coalescent model to all clusters containing >40 samples, giving 50 clusters (11 for the 614D variant and 39 for 614G).

Under this model, population size grows exponentially up to a transition time, whereupon it shrinks exponentially. Rates of growth and decline and the transition time can vary for each 614G and 614D cluster but a joint estimate for these are obtained using a hierarchical model (see methods). Among the 50 clusters, the 614G clusters tended to start later and persist longer than 614D (Figure S4), whilst 614D clusters tended to have slightly earlier transition times (614D mean = 25th March, 614G mean = 1st April). We do not detect any significant evidence for positive selection of the 614G variant using this model (Table 1), as uncertainty in estimated cluster growth rates was large (Figures 3 and S5). Growth rates for 614G clusters tended to be larger (posterior mean = 114 year^-1^, versus 93 year^-1^) as too were the decline rates of 614G clusters (posterior mean = −11 year^-1^, versus −9 year^-1^) but these differences were non-significant.

Further, we applied a non-parametric phylodynamic model that allows virus population size growth rate to vary over time according to a stochastic process. We applied this model independently to each of the clusters described above. We found that effective population size in the largest clusters tracks the progression of the epidemic in the UK and growth in most clusters is negative by early April 2020 (Figure S6 A-D). We then examined if the 614G variant explained variance in growth rates among phylogenetic clusters. The initial growth rate of each cluster was highly variable (Figure S6) and precision of the estimated rate was generally low. The Spike protein 614 polymorphism on its own explains very little variance in growth rates among clusters (weighted least squares R^2^=1%) and there was no significant difference in initial growth rates (median initial growth rate for 614D clusters = 117 year^-1^, versus 169 year^-1^for 614G clusters; Kruskal Wallis p = 0.13). This corresponds to an R_0_ of 3.1 (interquartile range, IQR: 2.7-3.5) for 614D clusters and 4.0 (IQR: 3.1-4.8) for 614G clusters, assuming a 6.5 day serial interval (Flaxman et al., 2020). The region of sample collection was not significantly associated with growth rates (weighted least squares, p=0.248). We did not observe a significant association between growth rates and the first detection date of a cluster (weighted least squares, p=0.62).

We next examined if there was a detectable difference in growth rates by combining information from the virus phylogeny and the empirical frequency of sampling of the 614G variant over time. We conducted a model-based phylodynamic analysis using 200 sequences sampled randomly from the London metropolitan area (see Phylodynamic Methods). A phylogeographic model specified the relationship between the London sequences and a random sample of 100 sequences from outside of London, thereby providing a mechanism to control for founder effects. Figure 3D shows the estimated frequency of 614G and 614D infections over time in London using this approach. We estimated that 614D was initially the most prevalent variant but that 614G overtook 614D in late March. A similar transition from 614D to 614G was observed in the empirical sampling frequencies, such that by the end of March samples from London are more than twice as likely to be the 614G variant. The phylogeographic model was fitted both with and without information about sampling frequency of 614G over time. Incorporating sampling information into the mode increases the estimated selection coefficient, from 0.10 (without sampling information) to 0.26 (95% CI: −0.01-0.58) (Table 1). It is important to note that all fitted trajectories predict that the log odds of sampling 614G increase even if the selection coefficient is zero and that this is not necessarily evidence of positive selection for 614G.

### Association of Spike 614 replacement with infection severity, outcome, and age

We investigated associations between the D614G polymorphism and virulence by linking virus genome sequence data with clinical data on patient outcomes. We studied two clinical outcome datasets: Dataset 1 – 9,782 614G and 2,533 614D associated genetic sequences collected by Public Health England between 3 February and 4 July, 2020 linked to patient outcome after 28 days post-diagnosis (death or recovery), and Dataset 2 – 1,670 (486 614D and 1,184 614G) genetic sequences collected by NHS Greater Glasgow & Clyde between 28th February and 30th June 2020 linked to records of clinical severity. In univariate analyses of dataset 1, we found that patients with the 614G variant show reduced odds of death, but this effect disappeared after controlling for other known risk factors for severe COVID19 outcomes (Table 2). Mortality closely tracks average age within our sample which varied greatly over time as testing priorities changed (Figure 3 E and F). We observed associations between time of sampling and genotype (later samples were more likely to have 614G) and later samples having higher odds of death and higher age. Odds of survival decrease for later samples, which may reflect prioritization of very severe cases for hospitalization and genetic sequencing as the epidemic peaked in March and April. For Dataset 2, clinical severity was recorded using an ordinal scale based on oxygen requirement (1. No respiratory support, 2: Supplemental oxygen, 3: Invasive or non-invasive ventilation or high flow nasal cannulae, 4: Death). The association between the D614G polymorphism and severity of disease was estimated with high uncertainty, but the posterior was centered close to zero indicating that a biologically relevant effect is unlikely (mean: 0.03; 95% CI: −0.80-0.84). Increasing age and male biological sex were both associated with a marked increase in clinical severity (Figure 4, Table S1). We found a correlation in infection severity between individuals infected with related viruses (mean standard deviation of the phylogenetic random effect: 0.26; 95% CI: 0.19-1.09). However, it is unclear to what extent this correlation represents genetic differences between viruses underlying infection outcomes as opposed to being an artefact of related viruses being spatially co-located, and thus infecting individuals with similar characteristics.

**Table 2:** Odds ratios (OR) of death within 28 days post diagnosis. Continuous variables were scaled (Z score) before regressing. Coefficients are in standardized units (Z-score). 95% confidence intervals are shown in parentheses.

| Predictor | OR | Adjusted OR | Coefficient |
| --- | --- | --- | --- |
| 614 G | 0.82 (0.74 - 0.90) | 1.09 (0.97 - 1.23) | 0.09 (-0.03 - 0.21) |
| Sex=Male |  | 2.15 (1.95 - 2.36) | 0.77 (0.67 - 0.86) |
| Age |  |  | 1.63 (1.56 - 1.70) |
| Time of sampling |  |  | -5.6(-6.68 - -4.62) |

**Figure 4.**
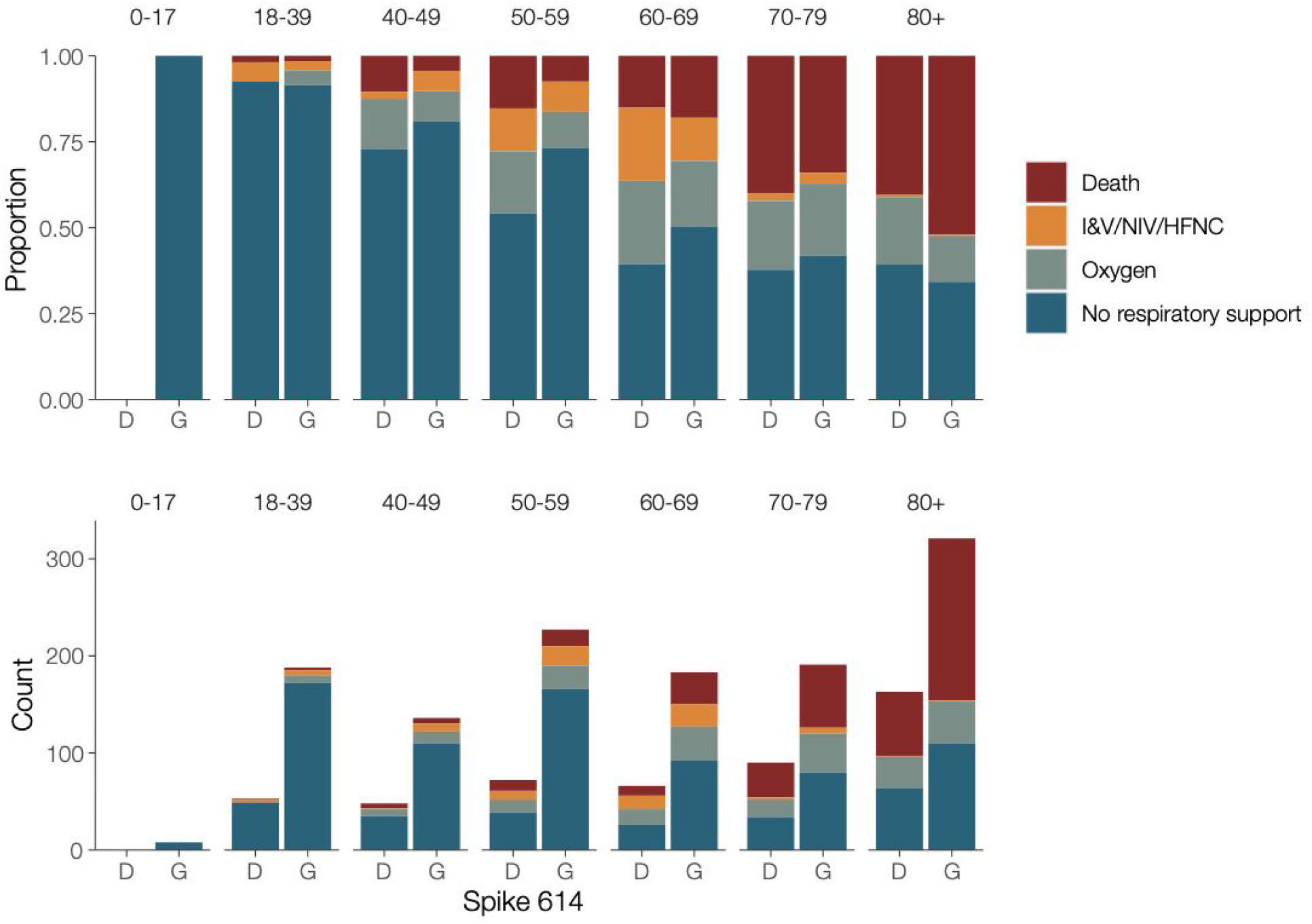
Clinical severity in patients in association with the D614G polymorphism and age. Clinical severity was measured on a 4 point ordinal scale based on requirement for respiratory support. Upper panel – proportion of outcomes by age, lower panel – absolute counts. I&V – intubation and ventilation; NIV – non-invasive ventilation; HFNC – high flow nasal cannulae; Oxygen – supplemental oxygen delivered by face mask or low-flow nasal cannulae.

We observed an association between age and genotype, with younger patients more likely to carry 614G viruses. We see this association despite the progressive aging of the patient cohort (Figure 3F) and concomitant increase in prevalence of 614G relative to 614D. We performed a multivariate analysis on the metadata of 27,038 sequences from across the UK (England, Wales, Scotland, and Northern Ireland) for the sample collection date, the age and sex of patients. A significant difference was found between the distribution of patient ages for 614G and 614D (Figure S8, Mann Whitney U test: p < 10^-13^). The median age is 5 years older among female carriers of 614D versus 614G and 4 years older among male carriers of 614D versus 614G. An association was also observed between sex and the presence of 614G or 614D (Figure S8, Chi-squared test: p < 10^-10^). Differences in the age distribution for each sex were also observed (Mann-Whitney U p<10^-8^ for 614D and p<10^-37^ for 614G). The probability of carrying 614G virus seems to decrease continuously with age (Figure S8). This is possibly due to an increased viral load in younger patients associated with 614G variants leading to higher detection rates.

As a proxy for viral load we studied 12,082 sequences with PCR cycle threshold (Ct) values from across the UK. Sequences with 14≤ Ct ≤ 40 were inspected for association with genotype and a very slight (<1 Ct step) but significant difference was observed with 614G associated with lower Ct (Figure S6, p <10^-6^, Mann-Whitney U test). As different test methods were used to obtain the Ct values across the dataset making a reliable comparison difficult, we carried out real-time quantitative viral load testing using a subset of 31 614D and 290 614G samples extracted on the same platform and analysed using the 2019-nCoV_N1 assay RT-qPCR assay. This again found a significant difference with 614G associated with higher viral load (Figure 3G, p=0.0151, Mann-Whitney U test).

### Other proximal residue replacements with potential relevance to Spike subunit function stability

Within the UK and global SARS-CoV-2 phylogenies there are multiple instances of the D614G mutation as well as reversions back to 614D. The existence of reversions implies that the 614D wild-type is still relatively fit within individual hosts. Within the UK, we also observe two phylogenetic clusters of another variant, 614N, the independent origins of which suggest that this variant is also transmissible. However, the effect of 614N on Spike subunit function remains to be determined.

We observed additional mutations at the residues immediately adjacent to Spike 614. The mutation 613H co-occurs with both 614G and 614D, possibly as a result of convergent evolution. V615I occurs on the background of 614D, whilst V615F co-occurs with 614G (Table 3). These replacements are associated with one or two UK clusters, showing evidence for their transmission within the UK. Variant 615I is largely constrained to Wales, where it is associated with a large phylogenetic cluster that has not been observed since mid-April. Experimental studies will be required to determine whether these mutations proximal to site 614 have similar effects to 614G or, when co-occurring, have compensatory or epistatic effects.

**Table 3:**
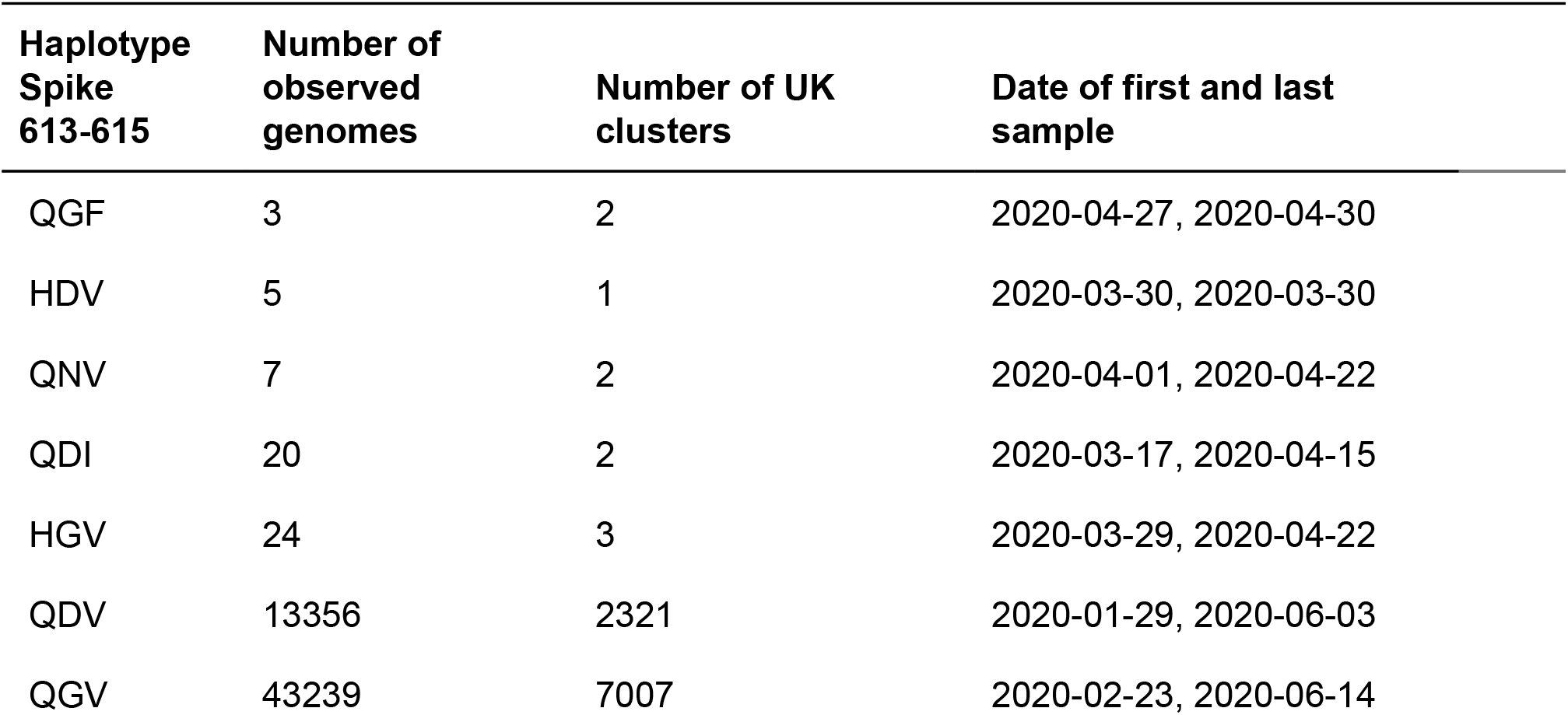
Circulating amino acid haplotypes found at residues 613-615 of the SARS-CoV-2 Spike protein. The ancestral haplotype is inferred to be QDV. The table reports the number of distinct UK clusters that the respective genomes are found in.

| Haplotype<br>Spike<br>613-615 | Number of<br>observed<br>genomes | Number of UK<br>clusters | Date of first and last<br>sample |
| --- | --- | --- | --- |
| QGF | 3 | 2 | 2020-04-27, 2020-04-30 |
| HDV | 5 | 1 | 2020-03-30, 2020-03-30 |
| QNV | 7 | 2 | 2020-04-01, 2020-04-22 |
| QDI | 20 | 2 | 2020-03-17, 2020-04-15 |
| HGV | 24 | 3 | 2020-03-29, 2020-04-22 |
| QDV | 13356 | 2321 | 2020-01-29, 2020-06-03 |
| QGV | 43239 | 7007 | 2020-02-23, 2020-06-14 |

## Discussion

The variant D614G has been shown to enhance the infectivity of pseudotyped lentiviruses carrying SARS-CoV-2 spike protein *in vitro* (Korber et al., 2020; Yurkovetskiy et al.). However differences in cell infectivity *in vitro* do not necessarily result in greater within-host infectivity, let alone increased transmissibility between hosts. The spread of a virus mutation is governed by demographic processes such as population growth, range expansion, founder effects and random genetic drift, as well as by potential positive selection if the mutation confers enhanced transmissibility. We used available data to indirectly evaluate the transmission fitness of the Spike 614G by using a very large dataset of patient samples and a range of inference approaches. Some methods suggested evidence for increased population growth rates of the 614G variant whilst others did not. Given the many factors that contribute to transmission dynamics, it is unsurprising the population-level values we have estimated are much less than the proportional increase in cell infectivity measured *in vitro*.

Estimating the epidemiological fitness of individual genetic variants during an emerging pandemic presents multiple challenges. The recent origin of SARS-CoV-2 combined with a relatively low rate of evolution means global viral genetic diversity is low and many methods for identifying positive selection will have low sensitivity. Evidence for positive selection at Spike position 614 and other sites has been suggested by statistical models based on the rate ratio of nonsynonymous to synonymous substitutions (Pond, 2020). However the detection of positive selection by such methods does not necessarily imply the mutation enhances transmissibility, and effects of individual mutations on transmissibility will generally be low (MacLean et al., 2020).

Convergent molecular evolution (resulting in homoplasies) can present an alternative sources of information about potentially beneficial virus mutations (ADD REF) However, such approaches lack sensitivity for the D614G as almost all circulating 614G genomes derive from a single ancestor (van Dorp et al., 2020). Our discovery of co-occurring mutations in neighbouring sites (615 and 613) and the D614N variant is suggestive of a more complex selective landscape in this region of the Spike protein than was first indicated. We also note that our analysis is limited by necessity to the comparison of co-circulating clusters that, in some cases, are characterised by mutations at sites other than 614, hence it is impossible to disentangle the selective effects of each individual mutation. One amino acid replacement is notable: RdRp P323L, occurred almost concurrently with D614G, and is in almost perfect linkage equilibrium with 614G (Pond, 2020).

We have drawn on two sources of information regarding the growth of the 614G variant, (i) the relative frequency of the 614G and 614D variants through time, and (ii) inferred differences in the genetic diversity and growth rate of 614G and 614D phylogenetic clusters in the UK (phylodynamics). Whilst the changing frequency of one variant in an exponentially-growing population can in theory indicate a difference in fitness, the rate at which 614G clusters were imported and discovered in the UK also changed through time, making direct comparisons of variant frequencies challenging. We attempted to control for this effect using phylogenetic analysis and by counting only samples derived from co-circulating clusters representing distinct introductions of SARS-CoV-2 into the UK. Separately, phylodynamic methods allow us to infer the growth and decline in effective population size of individual phylogenetic clusters, and we used this approach to compare the mean growth rates of 614G and 614D clusters. These phylodynamic estimates have high statistical uncertainty and do not consistently detect a significant difference in growth rate between the two variants. We observed, however, that 614G clusters tend to grow to a larger size than 614D clusters after controlling for time of introduction into the UK. This is consistent with a transmission advantage of 614G variants, but could also be the result of unknown confounders which increase the probability that 614G lineages will be sampled. Our data will be naturally biased towards samples that are easy to sequence, and we have observed a very slight but significant decrease in the RT-PCR Ct values of the 614G variant.

Several limitations of the data and analysis should be considered when interpreting our finding that the sampling frequency of variant 614G increased. We have applied classic population genetic models premised on contrasting the exponential growth rates of the 614G and 614D populations while controlling for founder effects, but in reality the SARS-CoV-2 epidemic is noisy and structured in ways not accounted for by this model. The frequency of 614G and 614D variants can change rapidly due to stochastic fluctuations, especially early in the epidemic. The sampling process is also inhomogeneous through time and sometimes reactive to short-term public health situations (e.g. nosocomial outbreaks) rather than being fully randomised and systematic. Most of the SARS-CoV-2 genome sequencing performed by centres in the UK is focused on symptomatic cases, often using diagnostic residual samples. As testing priorities change, and as cases in different segments of the population fluctuate, signals may emerge that are due to operational changes rather than shifts in virus biology.

Phylodynamic estimates of reproduction numbers are sensitive to the context of early spread of epidemic clusters which may have involved superspreading events (Endo et al., 2020). These events are highly variable and phylodynamic methods are inherently imprecise with poorly resolved phylogenies. The Spike 614 polymorphism explains little variance in the rate of spread of individual clusters, but incorporating additional information about the frequency of sampling 614G and 614D variants improved precision of phylodynamic estimates. Estimates of the reproduction number based on large clusters are not representative of the epidemic as a whole and may be larger on average.

SARS-CoV-2 case and infection fatality rates seem to vary widely among countries and through time. It is unclear to what degree this variation reflects estimation uncertainty, host population factors (such as the age structure of the population (Onder et al., 2020)) or virus genetic factors Here, we do not detect a difference in virulence between the two Spike 614 variants. By estimating mortality rates as opposed to rates of hospitalization or ICU care, our results complement those in (Korber et al., 2020) and are based on a substantially greater sample size. In addition, we did not find any association with clinical severity indicated by the requirement for oxygenation or respiratory support in a subset of 1670 patients. A significant association of 614G carriage with age may indicate minor differences in clinical outcome or frequency of symptomatic infection, which bears further study. The data is heavily skewed towards hospitalized cases, and therefore more severe disease, and so it is not possible to evaluate small differences in virulence that may be present in milder or asymptomatic infections. This is especially problematic for evaluating effects that may be confounded by age, as the proportion of infections that do not lead to symptoms is higher in younger individuals(Davies et al., 2020).

Our analysis emphasises that while laboratory experiments can identify changes in virus biology, their extrapolation to identify population level effects on transmission requires caution. In the case of D614G, a large increase in cellular infectivity results in a weak population-level signal that nonetheless produces a discernible effect on transmissibility. While we believe an effect on SARS-CoV-2 transmissibility caused by D614G is likely to be present, it is important to note that the estimation of the absolute size of this effect is uncertain and much harder to predict. Although the signal is difficult to detect, the unprecedented size and completeness of the UK dataset and associated metadata enables many potential biases within the data to be controlled for. This work is therefore demonstrative of the value of large-scale coordinated sequencing activities to understand a pandemic in real time.

This study shows that transmissibility of SARS-CoV-2 can change as the pandemic unfolds. Whether the current explosive epidemics across the world are to any degree being driven by D614G, or whether it is simply the beneficiary of being in the right place at the right time, it is now the dominant variant. Changes in the transmissibility of a circulating virus could have a major effect on pandemic planning and the effectiveness of pandemic response, and so it is critical that the parameters for models used for planning are based on the currently circulating virus. Work on vaccines, therapeutics and other interventions must allow for this but also keep in mind that reversions, and other mutations at the same or adjacent residues, will undoubtedly emerge in the future.

## Methods

### Sample collection and sequencing

We utilized data from the Coronavirus Disease 2019 (COVID-19) Genomics UK Consortium (CoG-UK)(The COVID-19 Genomics UK (COG-UK) consortium, 2020), a partnership of more than 18 academic, medical and public health research centres contributing sequencing and analysis capabilities. Sequence data was generated from a variety of protocols and platforms and were uploaded to a centralized environment for storage and analysis (MRC-CLIMB) (https://www.climb.ac.uk/KConnor et al., 2016). Data are uploaded with a standard set of clinical and demographic metadata and information about sequencing protocols and sample collection methods. Data undergo quality control and assembly and lineage assignment (Rambaut et al.). Data which complete quality control and assembly steps are released on a weekly basis. Sequence data are periodically shared through two open access databases, the European Bioinformatics Institute (Rodriguez-Tomé and Stoehr, 1996) and the Global Initiative on Sharing All Influenza Data(Shu and McCauley, 2017). We utilized 26,986 whole genome sequences contained in the June 19 release (https://www.coaconsortium.uk/data/) and for which the Spike 614 genotype could be determined and sample collection date was known.

### Phylogenetics and calculation of maximum parsimony clusters

Maximum likelihood (ML) phylogenetic trees were estimated separately using IQTree *v1.6.12* for major global lineages (Minh et al., 2020; Rambaut et al.). Phylogenies were rooted on a sample from the ancestral lineage. UK clusters were identified using parsimony-based ancestral state reconstruction (Fitch, 1977) with internal nodes classified as UK or non-UK. Most UK clusters are descended from polytomies with descendents in multiple countries, and reconstruction of ancestral states at such nodes is ambiguous. In such cases the polytomy node was assigned the same state as it’s ancestor. We consider two extremes of the maximum parsimony method for reconstructing ancestral states at bifurcating nodes: Accelerated transition (AT) which favours transitions to the UK as close to the root of the tree as possible, and delayed transition (DT) which favours transition to the UK as far from the root as possible. Unless otherwise stated, results are based on DT clusters which are more likely to represent transmission within the UK.

### Statistical analyses

Size of clusters was evaluated using log-linear multivariate regression. Effect of genotype on phylodynamic growth rates was estimated using multivariate weighted regression. Regression weights are inversely proportional to precision of estimated growth rates. Univariate comparisons used the Kruskal Wallis test. Kernel density estimation of sample time distributions used gaussian kernels and a bandwidth of 2 days. Statistical models were implemented in R 3.6.3.

### Logistic growth model

According to this model the number of infected with the Spike 614D variant grows exponentially at a rate *r* and the number with the Spike 614G variant grows exponential at rate 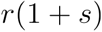. If *N_X_* is the number infected initially with variant *X*, the proportion of the population with Spike 614G at time *t* is

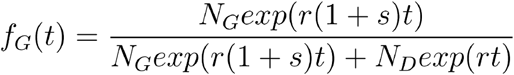

This model can be fitted to a sequence of sample times 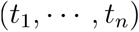 with Spike 614 genotypes 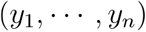 by maximum likelihood. The objective function is

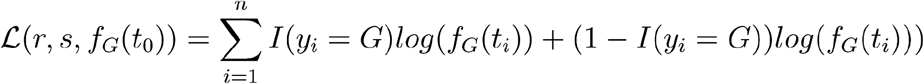

Formally, fitting this model is equivalent to logistic regression of genotype on time where the coefficient corresponds to the compound parameter 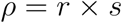. Deriving the selection coefficient therefore requires additional information about the growth rate *r*. For the model fitted to data during the exponential growth phase, we considered a range of plausible values for this rate corresponding to a reproduction number in the range 2.0-3.5 and a serial interval of 6.5 days (Flaxman et al., 2020). For the model fitted to data during the decline phase, we considered a rate corresponding to a generation time between 3 and 8 days. The final confidence interval is based on these ranges as well as the confidence interval of *ρ* computed using profile likelihood.

### Phylodynamic analysis of cluster growth rates with parametric coalescent model

We used a two-epoch coalescent model to estimate a period of exponential growth followed by an independently estimated period of exponential decline. Note that although we refer to growth and decline, the growth rates for both epochs can take either positive or negative values. The transition time from growth to decline was estimated independently for each cluster using a normal prior with a mean of the 23rd March 2020 (2020.2254), the date of ‘lockdown’ in the UK, and a standard deviation of two weeks. The data consisted of delayed transition clusters of more than 40 sequences as of the 19th June 2020.

A normal hyperprior is specified for cluster growth/decline rates for each genotype and the mean and precision of the hyperprior are estimated. The posterior mean growth/decline rates for each genotype are estimated along with the growth/decline rate for each cluster individually. Posterior growth rates within each genotype are therefore correlated. The prior for the mean growth rate is Normal(0,100/year) and the prior of the precision parameter is Gamma(1,0.001).

We compute the selection coefficient from growth rates with the formula 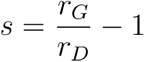 where *r* is the mean growth rate for each group of clusters.

The model was implemented in BEAST v1.10.5(Suchard et al., 2018). Four independent chains of 100m states were run for each variant, with 10% removed from each chain to account for burn-in. Convergence was assessed using Tracer(Rambaut et al., 2018) prior to further analysis. The HKY model was used to model nucleotide evolution(Hasegawa et al., 1985), and, following Duchene et al.(Duchene et al., 2020), the evolutionary clock rate was fixed at 0.001 substitutions per site per year. Other priors used are described in table S2.

### Phylodynamic analysis of cluster growth rates with non-parametric coalescent model

Rooted and dated phylogenies were estimated by randomly resolving polytomies in the ML trees described above using *ape 5.3*(*Paradis et al, 2004*) and *treedater* 0.5.1 (Volz and Frost, 2017). The mean clock rate of evolution was constrained to (0.00075,0.0015). Branch lengths were smoothed by enforcing a minimum number of substitutions per site on each branch and by sampling from the distribution estimated by *treedater*. This was carried out 20 times for each UK lineage. Growth rates were estimated using *skygrowth* 0.3.1 (Volz and Didelot, 2018) using Markov chain Monte Carlo (MCMC) and 1 million iterations for each time tree and using an Exponential(10^-4^) prior for the smoothing parameter. The final results were produced by averaging across 20 time trees estimated for each cluster. Code to reproduce this analysis is available at https://ait.io/JJklM and an interactive dashboard showing growth and decline of UK lineages can be viewed at https://shinv.dide.imperial.ac.uk/s614LineaaesUK/.

### Model-based phylodynamic analysis

We applied a susceptible-exposed-infectious-recovered (SEIR) model(Diekmann and Heesterbeek, 2000) for the SARS-CoV-2 epidemic in London linked to an international reservoir. The SEIR model assumed a 6.5 day serial interval. The estimated parameters included the initial number infected, the susceptible population size, and the reproduction number. The model included bidirectional migration to the region outside of London (both within the UK and internationally) at a constant rate per lineage. Evolution outside of London was modelled using an exponential growth coalescent. Additional estimated parameters include the migration rate, and the size and rate parameters for the exponential growth coalescent. This model was implemented in the BEAST2 PhyDyn package(Bouckaert et al.; Volz and Siveroni, 2018) and is available here: https://ait.io/JJUZv. The phylogenetic tree was co-estimated with epidemiological parameters. In order to make results comparable between 614D and 614G lineages, the molecular clock rate of evolution was fixed at a value estimated using all data in *treedater* 0.5.1. Nucleotide evolution was modeled as a strict clock HKY process (Hasegawa et al., 1985). To fit the model we ran 20 MCMC chains for 20 million iterations, each using 4 coupled MCMC chains (Müller and Bouckaert, 2020). Bespoke algorithms were used to exclude chains which failed to sample the target posterior. We used identical uninformative Lognormal(mean log=0, sd log = 1) priors for the reproduction number in 614G and 614D lineages.

The model was fitted to 614G and 614D sequence data separately before being combined for joint inference with the sample frequency data. This is carried out using a sampling-importance-resampling strategy(Smith and Gelfand, 1992). We sampled parameters from the posterior estimated from genetic data uniformly and computed importance weights using a sequential Bernoulli likelihood based on the estimated frequency of 614G and 614D overtime. Parameters resampled 1 million times with these weights yield our final estimate of the posterior.

The selection coefficient given a ratio of reproduction numbers is computed as follows:

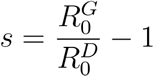

### Analysis of severity of patient outcomes

We aggregated data from 1670 patients presenting with COVID-19 from NHS records and combined it with the genome sequence of the virus infecting them. We used a phylogenetic generalised additive model to investigate the viral D614G polymorphism and association with severity of the infection.

To control for the effect of other mutations in the genome, we generated a time tree of the virus genomes from Scotland using an HKY + Γ nucleotide model excluding the nucleotide position underlying the D614G mutation. We estimated the tree using IQ-TREE 2 v. 2.0.6 (Minh et al., 2020). We masked the nucleotide causing the D614G mutation, as well as all mutations recommended by De Maio et al. as of 22/7/2020 (https://virological.Org/t/issues-with-sars-cov-2-sequencing-data/473/13). We included the first sequenced genome of SARS-CoV-2 from China (Wu et al., 2020) as an outgroup to root the tree.

We coded the severity of infection as four levels: 1) No respiratory support, 2) Supplemental oxygen, 3) Invasive or non-invasive ventilation or high flow nasal cannulae, 4) Death. We modified the WHO ordinal scale to these 4 points to avoid using hospitalisation as a criterion of severity because 1) many patients in nursing homes had severe infection but were not admitted to hospital, and 2) early in the outbreak, all cases were hospitalised irrespective of the severity of their infection. Our model included the presence of the D614G mutation and the biological sex of the patient as categorical predictors, as well as age and the time since the first case in the dataset as non-linear predictors. We include the time in days since the first case in the dataset to control for changes in treatment practice across the course of the epidemic. We mean-centred age and time in days and modelled their nonlinearities using penalised regression splines with a maximum of 30 knots. If a case was associated with a cluster of cases, for instance in a hospital ward or nursing home, this was included as a random effect with each cluster getting its own level. We gave any cases not associated with clusters their own unique level. Finally, to account for correlations driven by genome similarity that are not due to the D614G mutation, we generated a variance-covariance matrix (scaled to a correlation matrix) from the phylogeny described above (after dropping all tips corresponding to genomes not in the dataset) using the *ape* package v. 5.3 (Paradis and Schliep, 2019) and included that as a random effect in the model. We modelled the ordinal nature of the data using a cumulative model that assumes multiple thresholds corresponding to each severity level on the logit scale.

The model was fit in a Bayesian framework using Hamiltonian Monte Carlo in the R package *brms* v. 2.13.5 (Bürkner, 2018), a front-end for *rstan* v. 2.21.2 (Stan Development Team, 2020). The model had no divergent transitions, Gelman-Rubin values less than 1.01 and both bulk and effective sample sizes of greater than 950 for all parameters. Shortest probability intervals for reporting were generated by the R package *SPIn* v. 1.1 (Liu et al. 2015).

Priors: We used weakly informative priors to constrain the model to sensible values on the link scale, but not rule out any reasonable values. All thresholds for the dividing lines between severity levels were given t-distribution (mean = 0, scale = 2.5, df = 3) priors and all fixed effects were given Gaussian (mean = 0, standard deviation = 2.5) priors. The standard deviations for the random effects and penalised splines were given Exponential (lambda = 0.4) priors, corresponding to a prior mean of the standard deviation of 2.5, the same as the fixed effects.

### Clinical sample quantitative PCR

All samples were tested in duplicate using the 2019-nCoV_N1 assay RT-qPCR assay (https://www.fda.gov/media/134922/download); primers and probe were obtained ready-mixed from IDT (Leuven, Belgium). PCRs were performed in a final volume of 20 μl and included NEB Luna Universal Probe One-Step Reaction Mix and Enzyme Mix (New England Biolabs, Herts, UK), primers and probe at 500 nM and 127.5 nM, respectively, and 5 μl of RNA sample. No template controls were included after every seventh sample. Six ten-fold dilutions of SARS-CoV-2 RNA standards were tested in duplicate in each assay; standards were calibrated using a plasmid containing the N sequence that had been quantified using droplet digital PCR. Thermal cycling was performed on an Applied Biosystems™ 7500 Fast PCR instrument running SDS software v2.3 (ThermoFisher Scientific) using the following conditions: 55oC for 10 minutes and 95oC for 1 minute followed by 45 cycles of 95oC for 10 s and 58oC for 1 minute. Assays were repeated if the reaction efficiency was <90% or the R2 value of the standard curve was ≤0.998. Where possible, testing of samples was repeated if the %CV of the duplicates was <10%. Three samples were not tested in duplicate because of insufficient RNA. Two samples had Cq values that were below the top SARS-CoV-2 RNA standard in the assay. Duplicate PCRs from four samples had %CVs >10 (range 10.19 to 17.06).

## Data Availability

All sequence data and metadata used in this work is shared via:
The COG-UK website : https://www.cogconsortium.uk/data/ 
GISAID: https://www.gisaid.org/
and the ENA as part of Bioproject PRJEB37886: https://www.ebi.ac.uk/ena/data/view/PRJEB37886

https://www.ebi.ac.uk/ena/data/view/PRJEB37886

## Acknowledgements

We thank all partners and contributors to the COG-UK consortium who are listed at https://www.coaconsortium.uk/about/. We also acknowledge the important work of SARS-CoV-2 genome data producers globally contributing sequence data to the GISAID database, and particularly acknowledge the groups who have generated data used by this project, listed in Table S4. EV acknowledges the MRC Centre for Global Infectious Disease Analysis MR/R015600/1. VH was supported by the Biotechnology and Biological Sciences Research Council (BBSRC) [grant number BB/M010996/1], JTM, RMC, NJL and AR acknowledge the support of the Wellcome Trust (Collaborators Award 206298/Z/17/Z – ARTIC network). AR is supported by the European Research Council (grant agreement no. 725422 – ReservoirDOCS). DLR, ASF and ECT are supported by the MRC (MC_UU_1201412). JS was supported by the Biotechnology and Biological Sciences Research Council-funded South West Biosciences Doctoral Training Partnership [training grant reference BB/M009122/1], TRC and NJL acknowledge support from the MRC which funded computational resources used by the project [grant reference MR/L015080/1]. TRC acknowledges funding as part of the BBSRC Institute Strategic Programme Microbes in the Food Chain BB/R012504/1 and its constituent projects BBS/E/F/000PR10348 and BBS/E/F/000PR10352], AP and TRC acknowledge support from Supercomputing Wales, which is part-funded by the European Regional Development Fund (ERDF) via Welsh Government. The project was also supported by specific funding from Welsh Government which provided funds for the sequencing of a subset of the Welsh samples used in this study, via Genomics Partnership Wales.

## Supporting figures and tables

**Figure S1.**
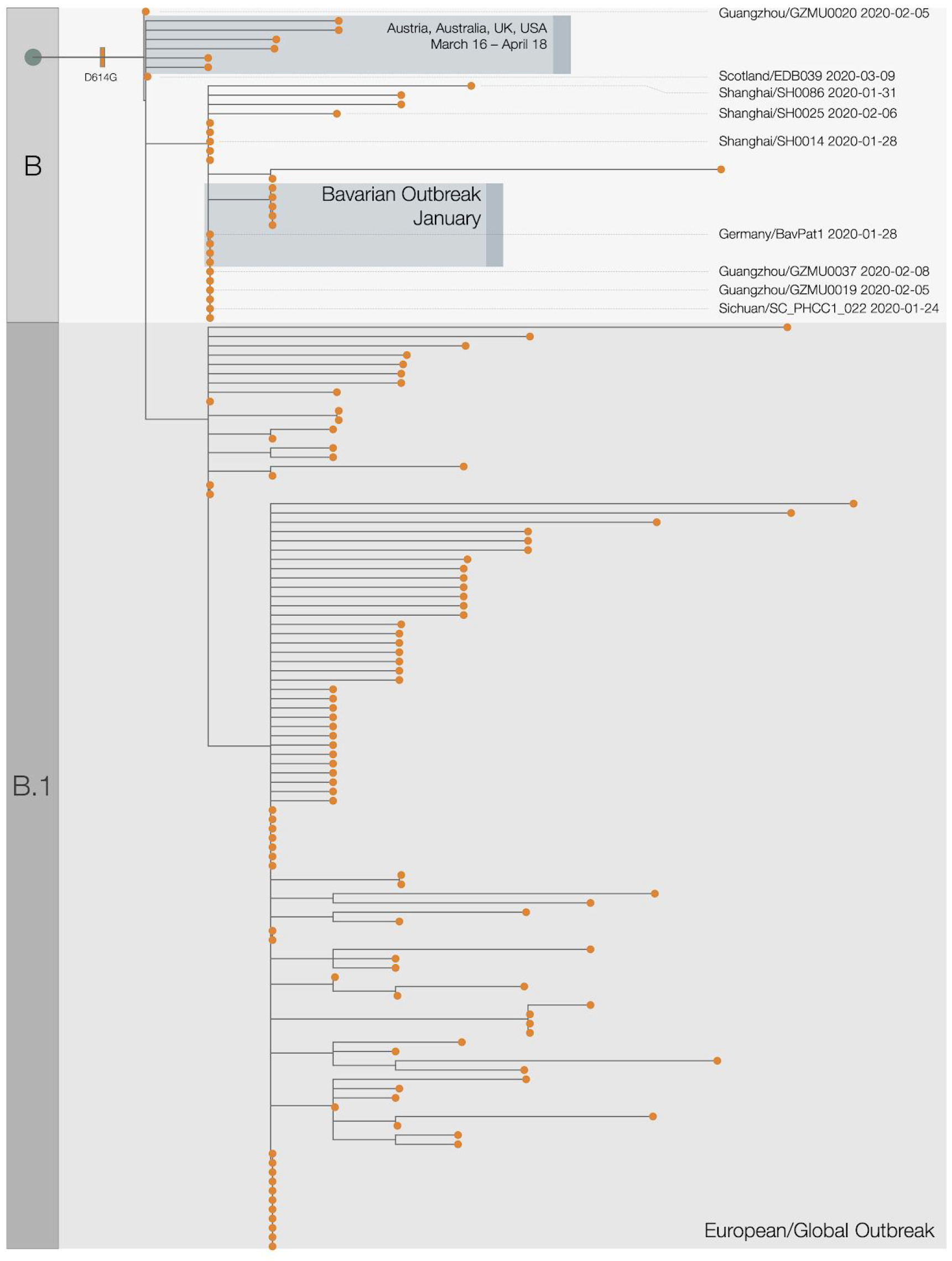
Expanded phylogenetic tree showing the early stages of emergence of D614G into Europe from China. Acknowledgments and details for highlighted genome sequences are given in Table S3.

**Figure S2:**
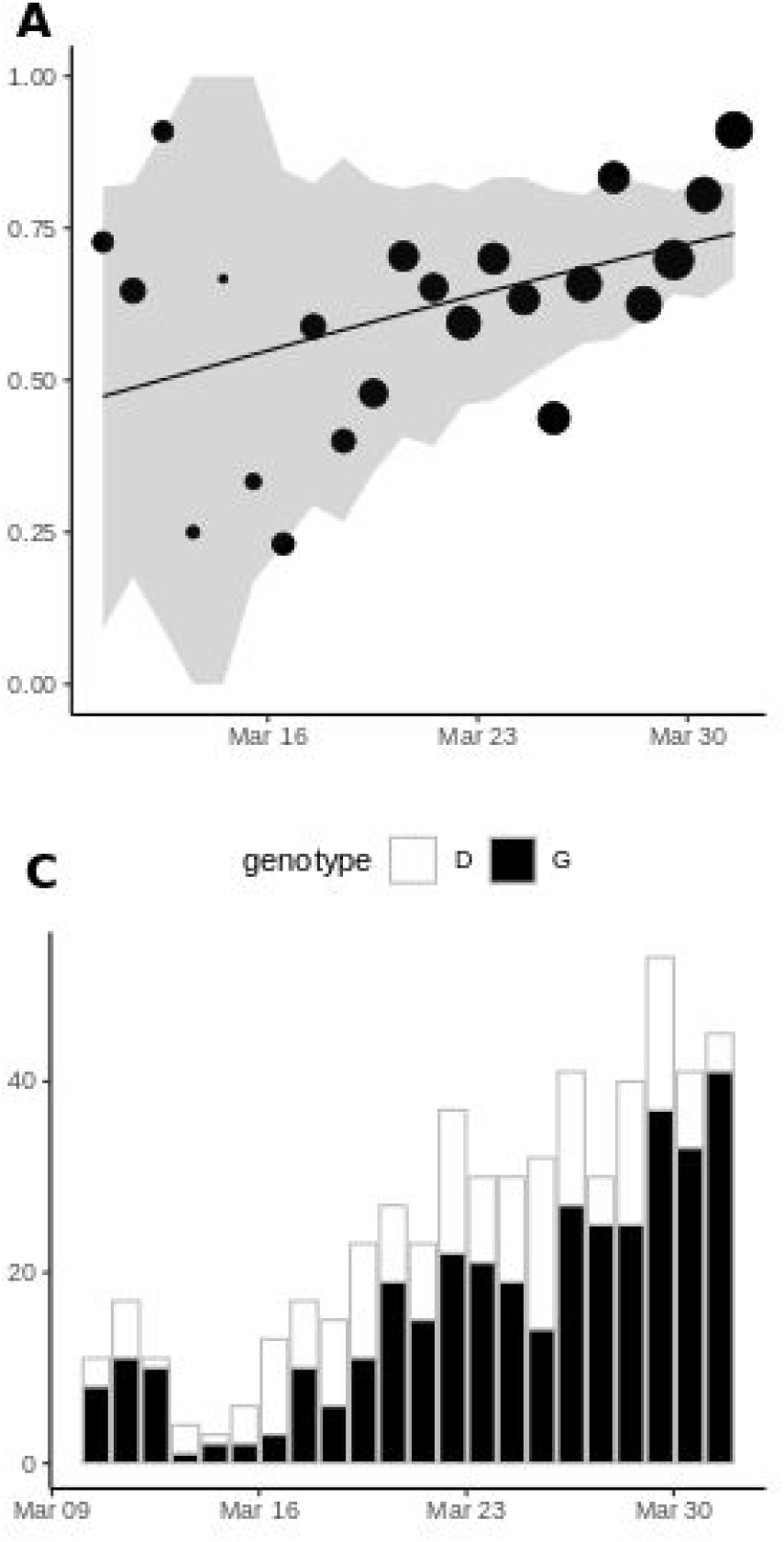
Frequency of sampling Spike 614G over time (A) and numbers of Spike 614G and Spike 614D samples over time(B). The size of points represents the number of samples collected on each day. The line and shaded region showed the MLE and confidence interval fit of the logistic growth model.

**Figure S3:**
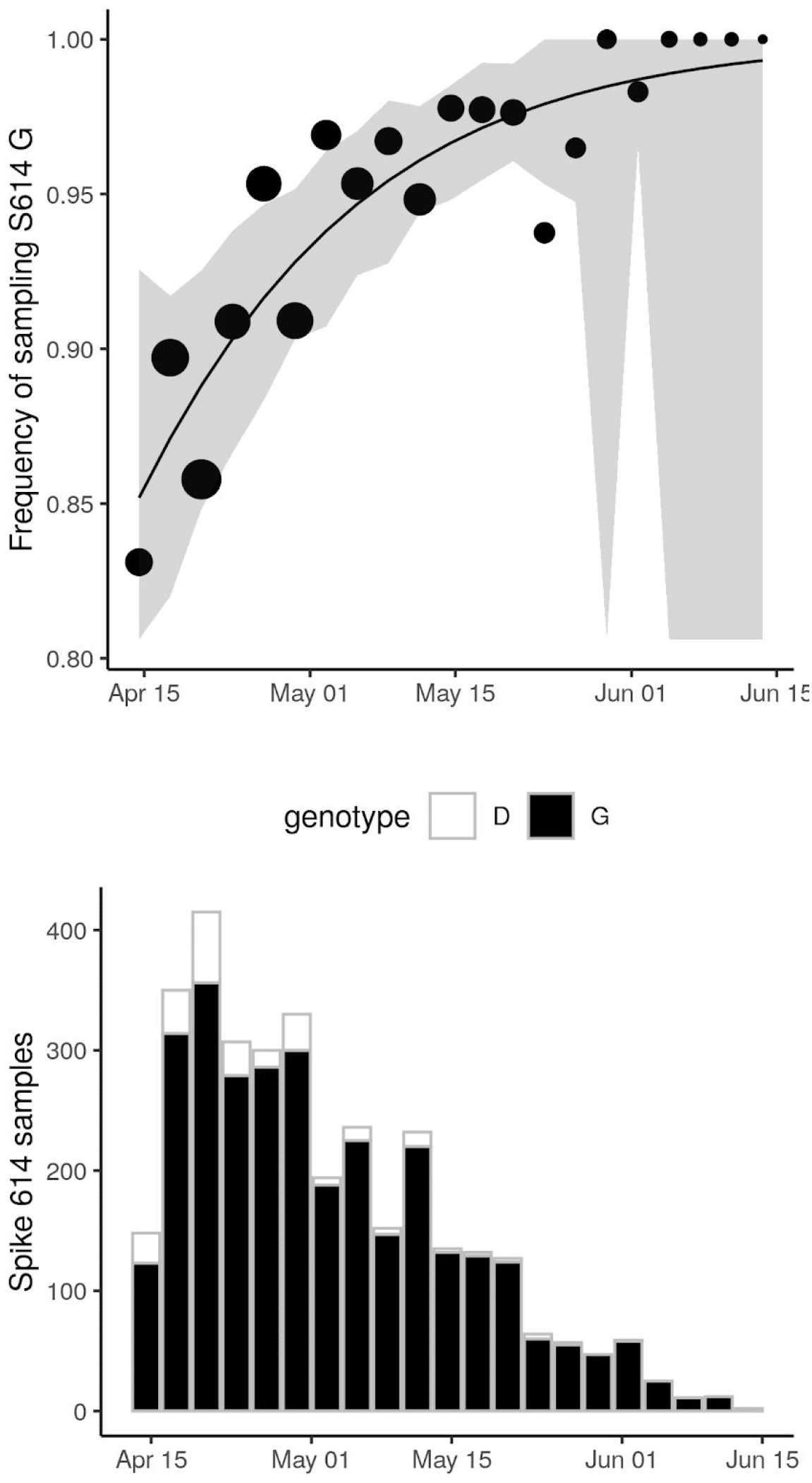
Frequency of sampling Spike 614G after April 15 and numbers of Spike 614G and Spike 614D samples overtime using 37 DT clusters detected before March 31, 2020. The size of points represents the number of samples collected on each day. The line and shaded region showed the MLE and confidence interval fit of the logistic growth model.

**Figure S4:**
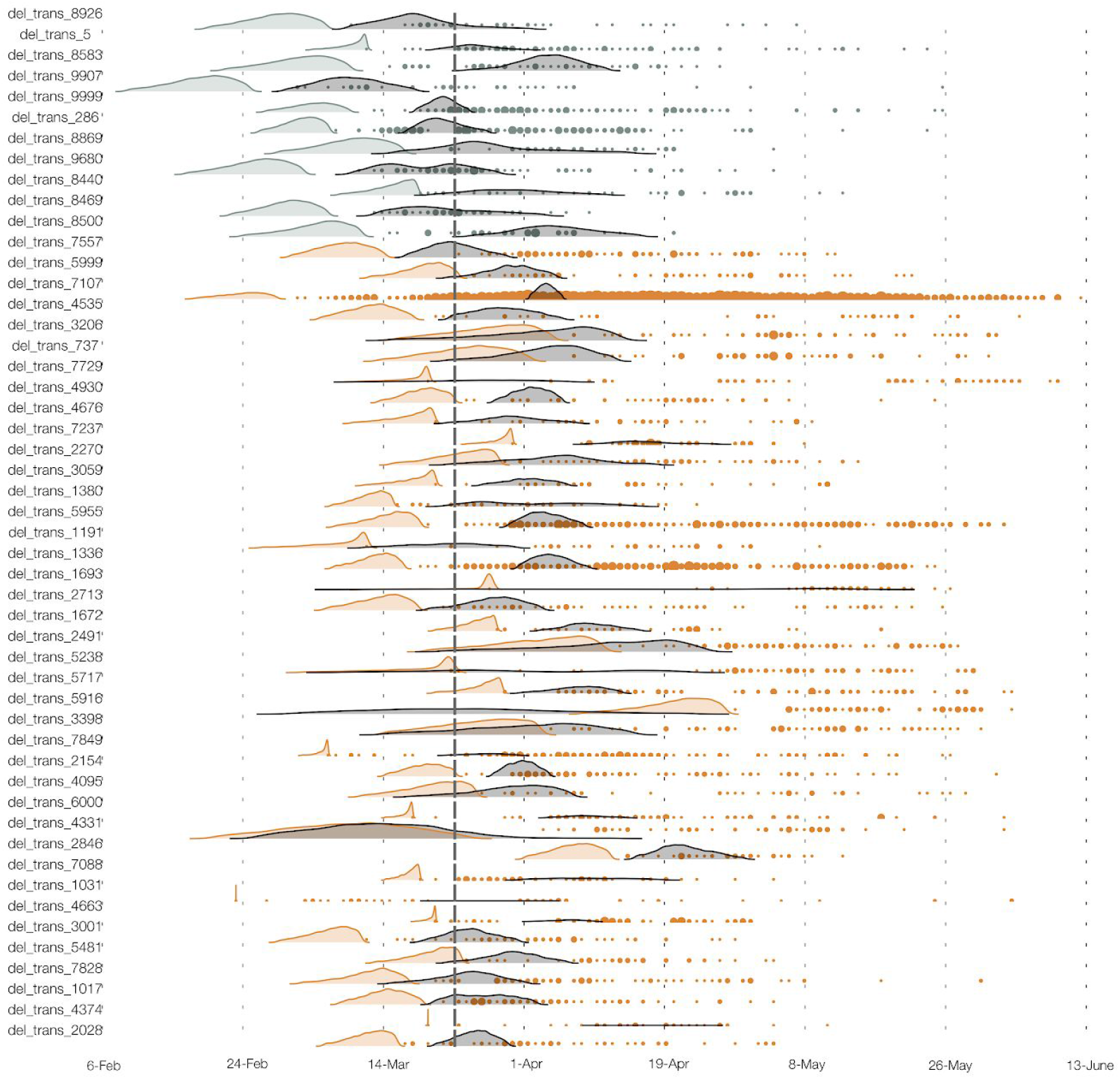
The estimated TMRCA for each of 50 UK clusters (shaded density) and time of each sequence sampled (points). Brown and grey respectively indicate Spike 614G and 614D clusters.

**Figure S5:**
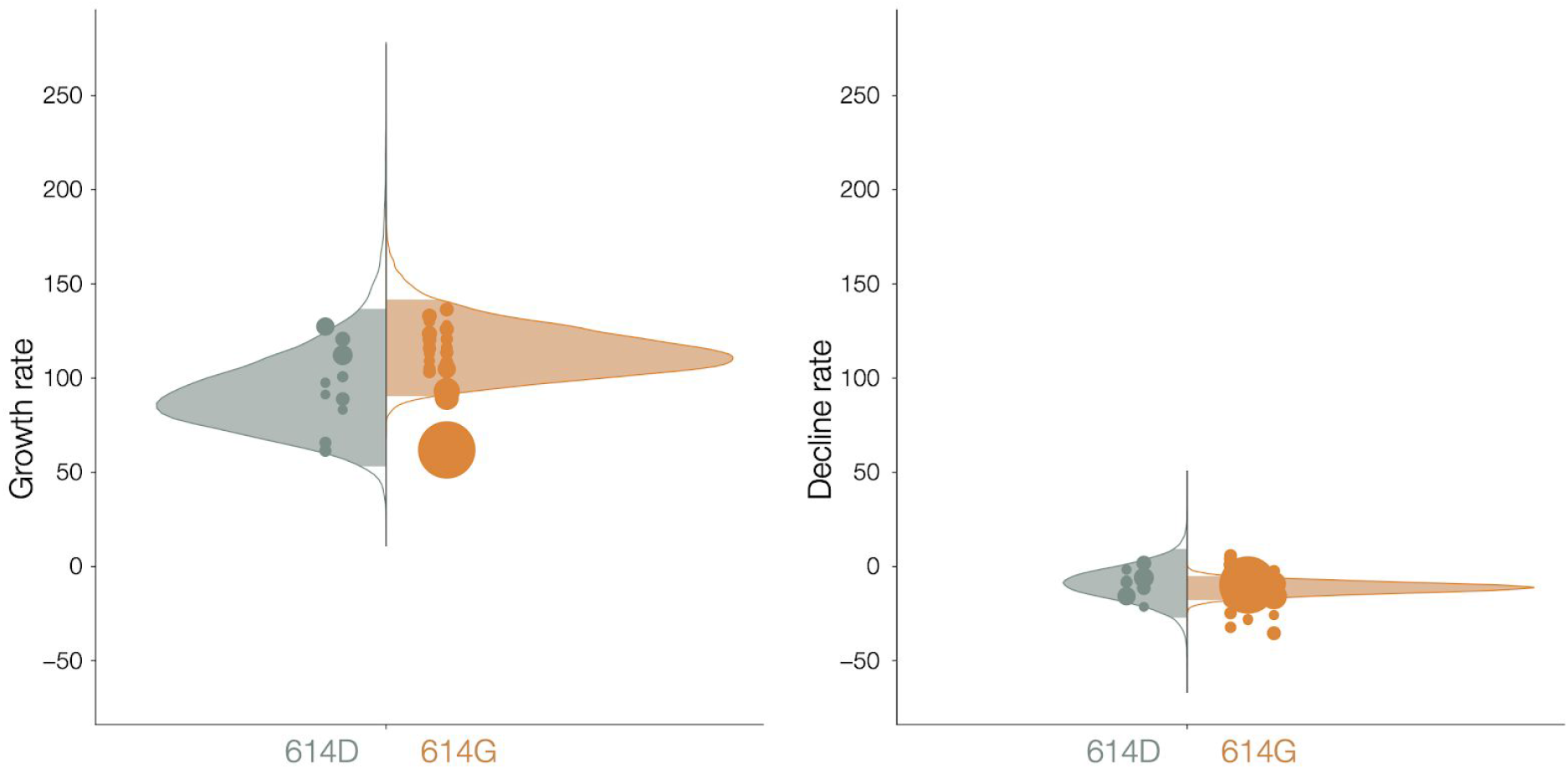
Distribution of exponential growth rates (left) and rates of decline(right) for Spike 614G (brown) and 614D (grey) in units of 1/year. Solid areas span the 95% credible interval. Points indicate the rates estimated for specific clusters, and are sized by the number of sequences in that cluster.

**Figure S6:**
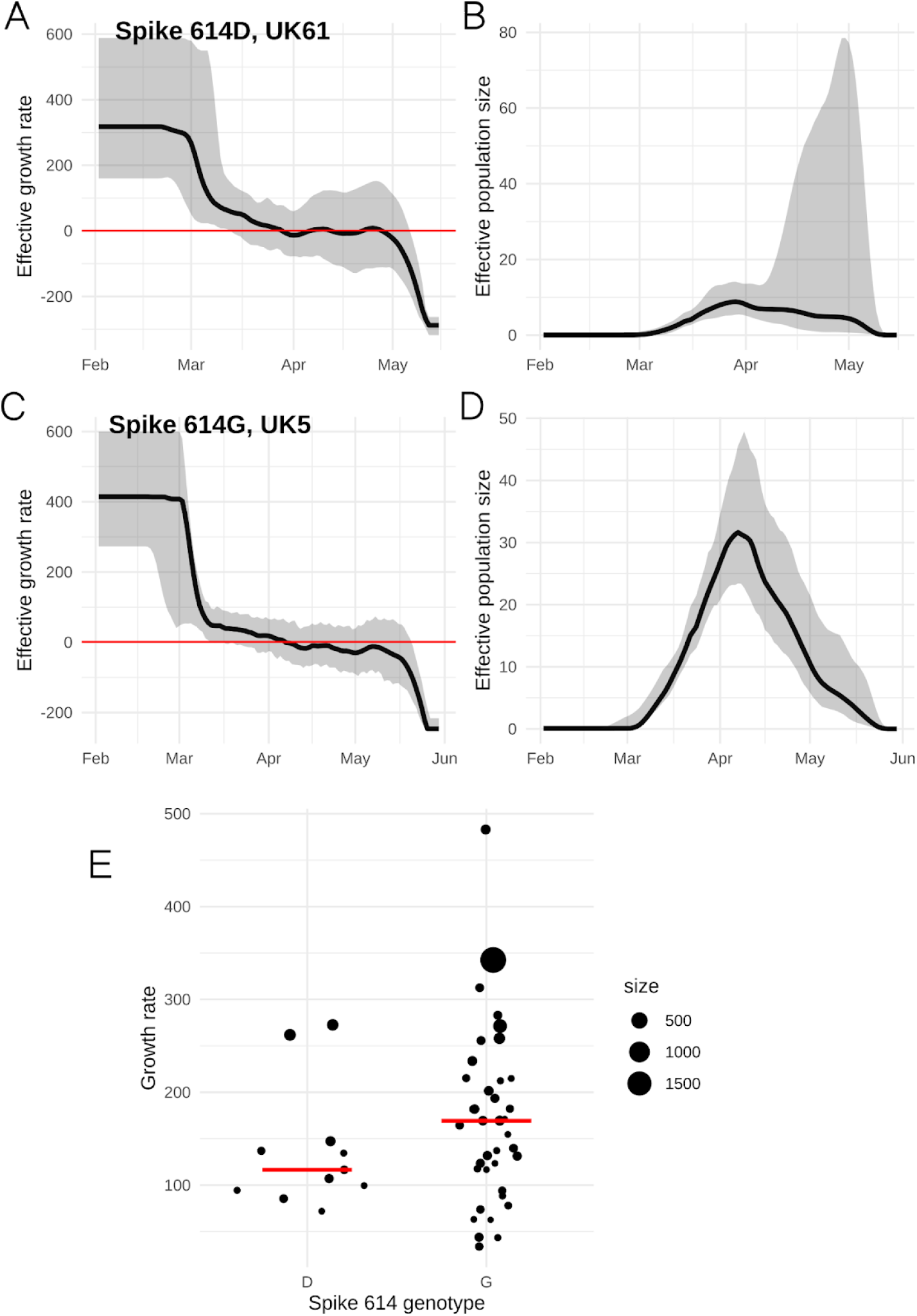
Estimated growth rates (A and C) and effective population size (B and D) for the two largest clusters with genotypes Spike 614D/G. The growth rate at the beginning of the time axis (Feb 1, 2020) is shown in panel E and provides a data point for the statistical comparisons between clusters. The size of points corresponds to the number of samples in each cluster.

**Figure S7.**
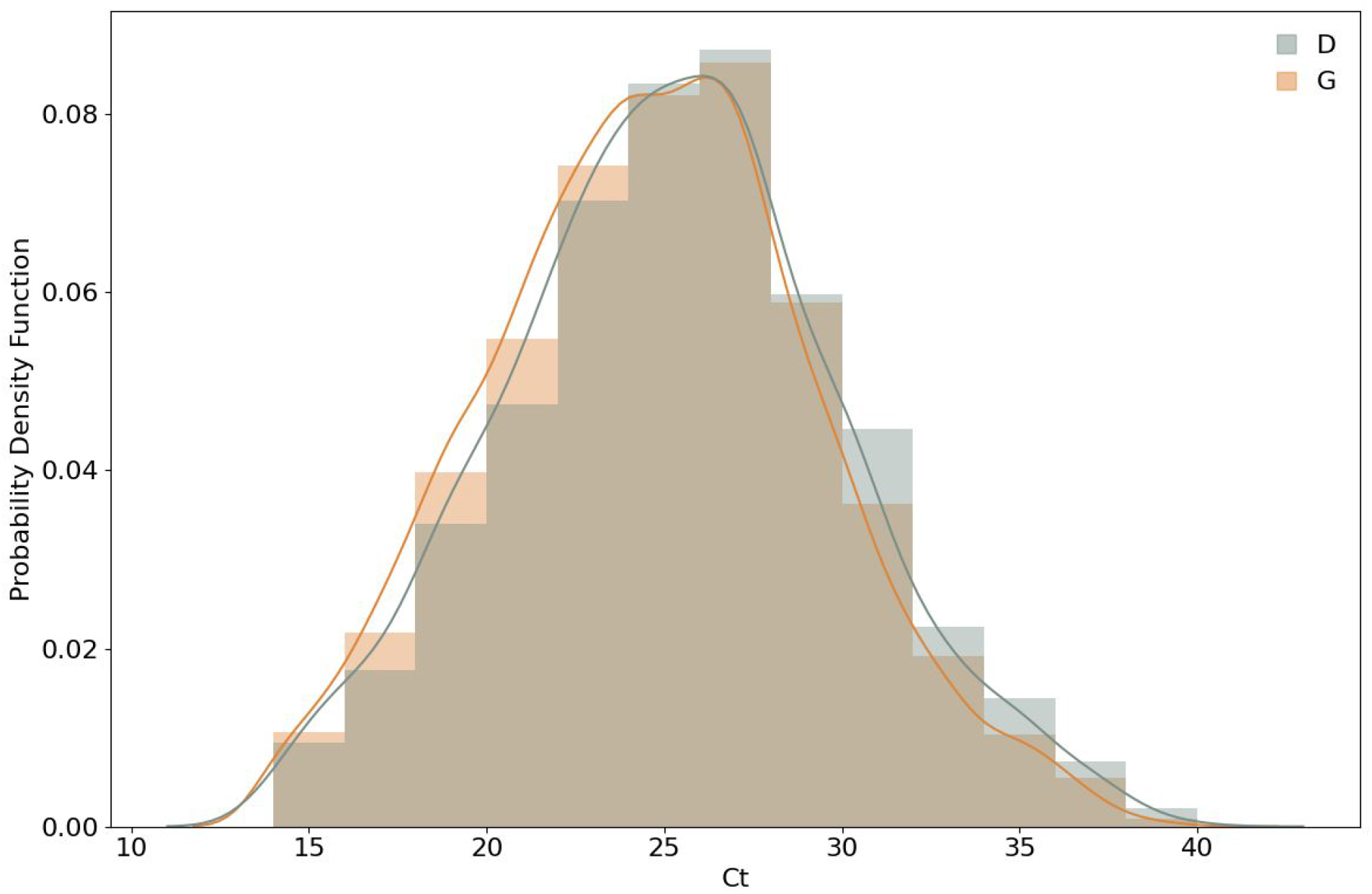
Frequency histograms of PCR cycle threshold (Ct) for D/G variants overlaid with kernel density estimates. Samples where the amino acid at position 614 was not recorded and samples with a Ct value of less than 14 or greater than 40 were excluded.

**Figure S8:**
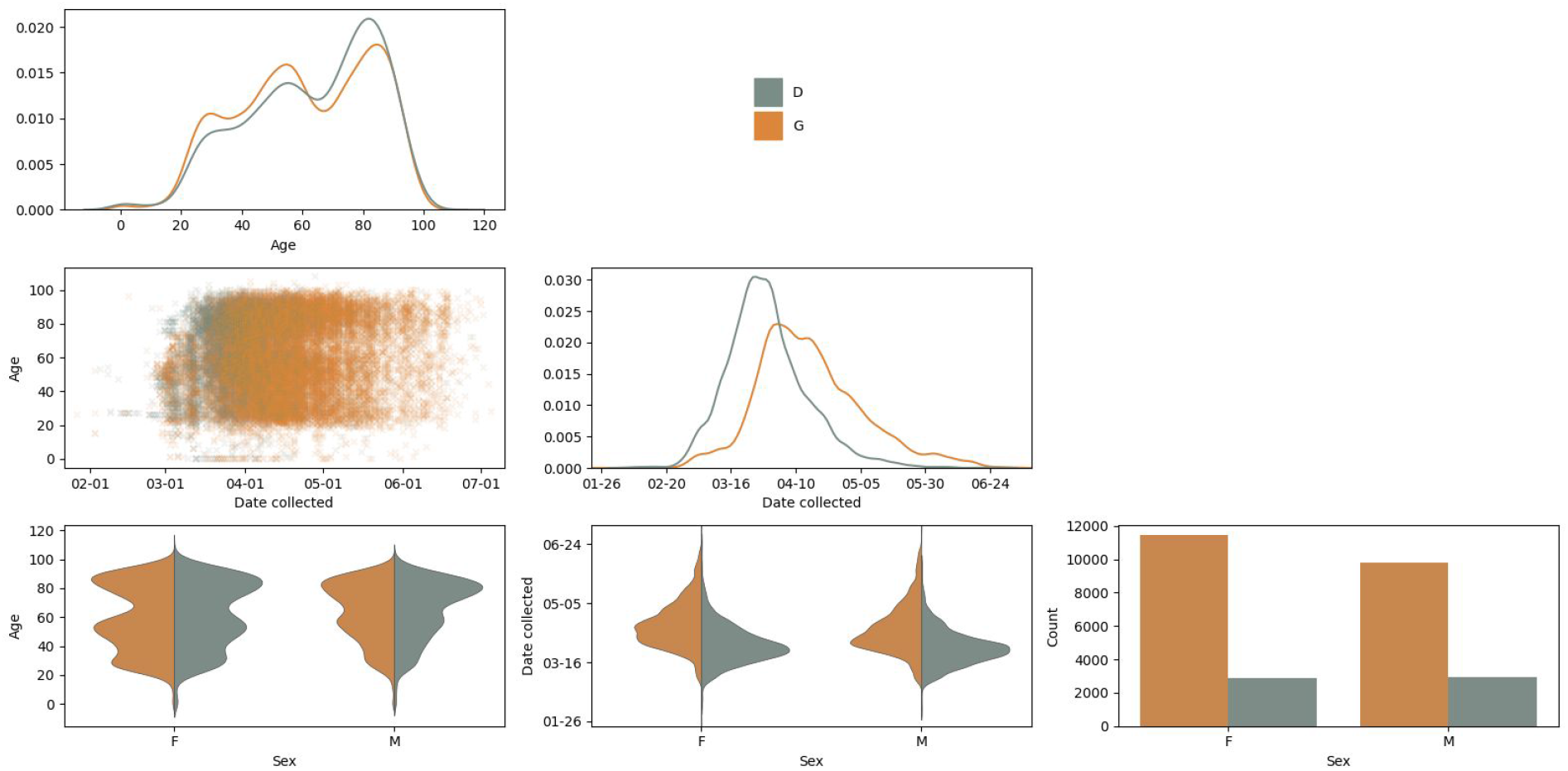
Probability of observing Spike 614G virus in patients grouped by age and sex. Collected pairwise plots are based on a UK-wide (England, Wales, Scotland, and Northern Ireland) multivariate dataset for the sample collection date, and the age and sex of the patient. Kernel Density Estimation (KDE) and count plots are on the diagonal.

**Table S1.**
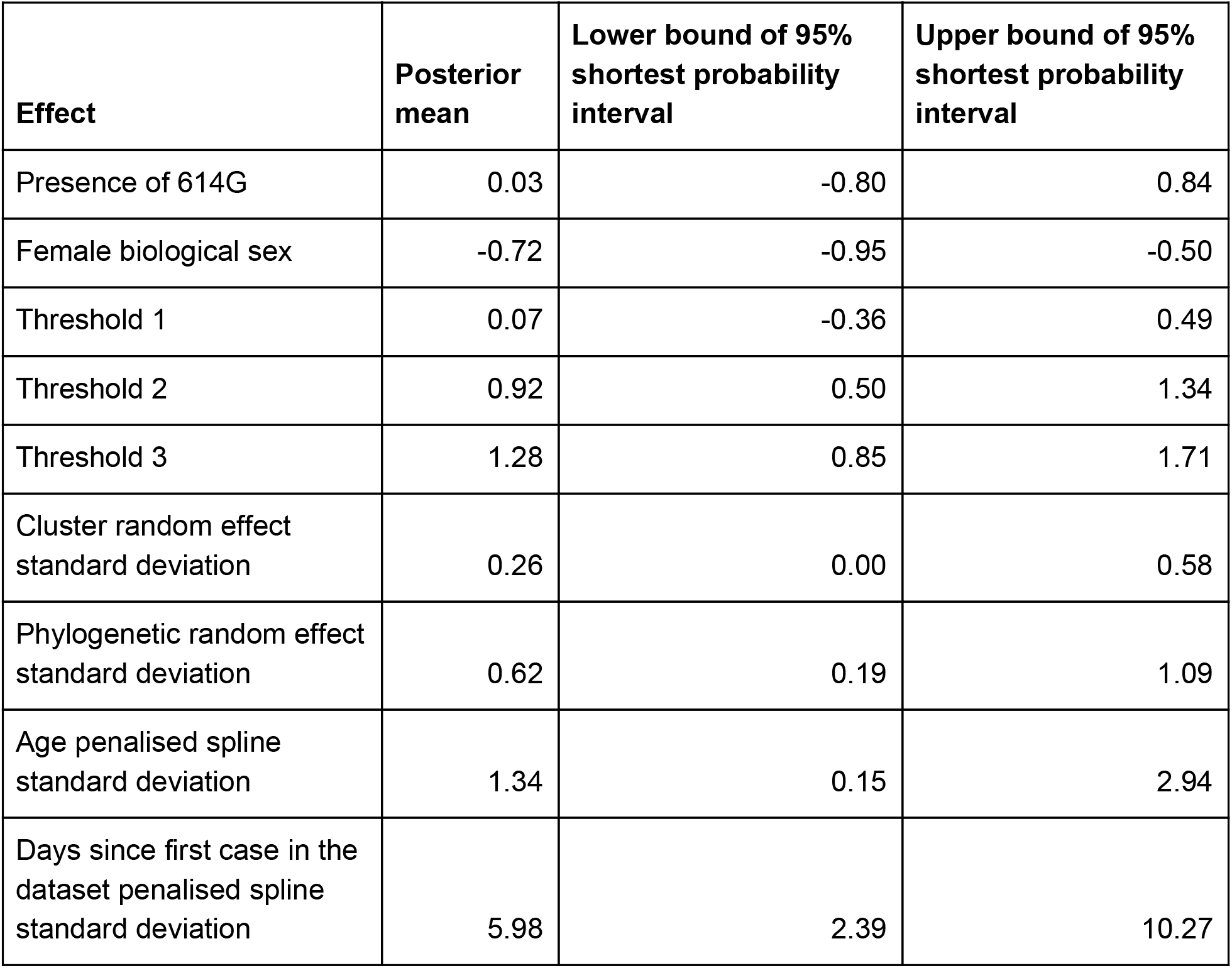
Posterior values for regression coefficients and standard deviations for the severity analysis of Dataset 2

**Table S2.**
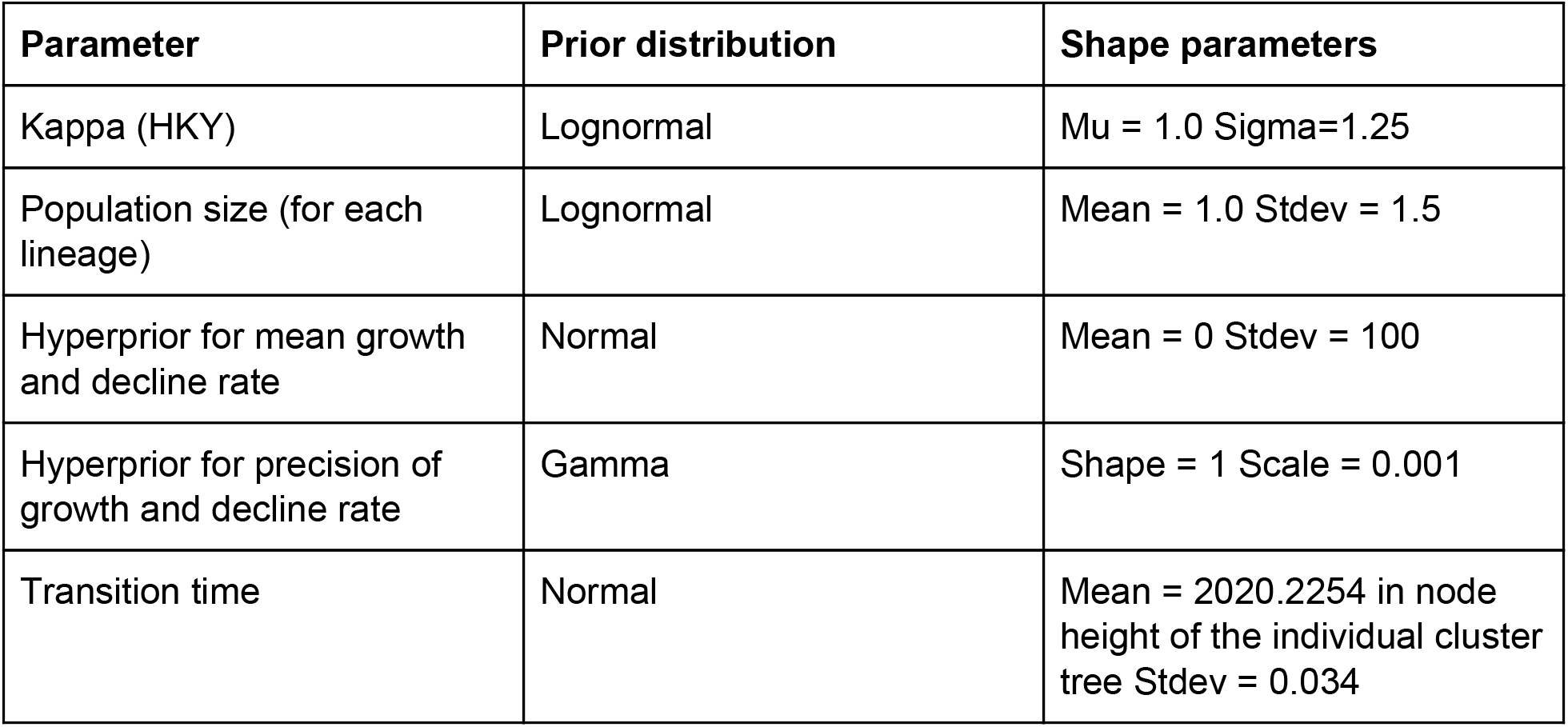
Prior distributions used in the BEAST analysis (see Main text).

**Table S3.**
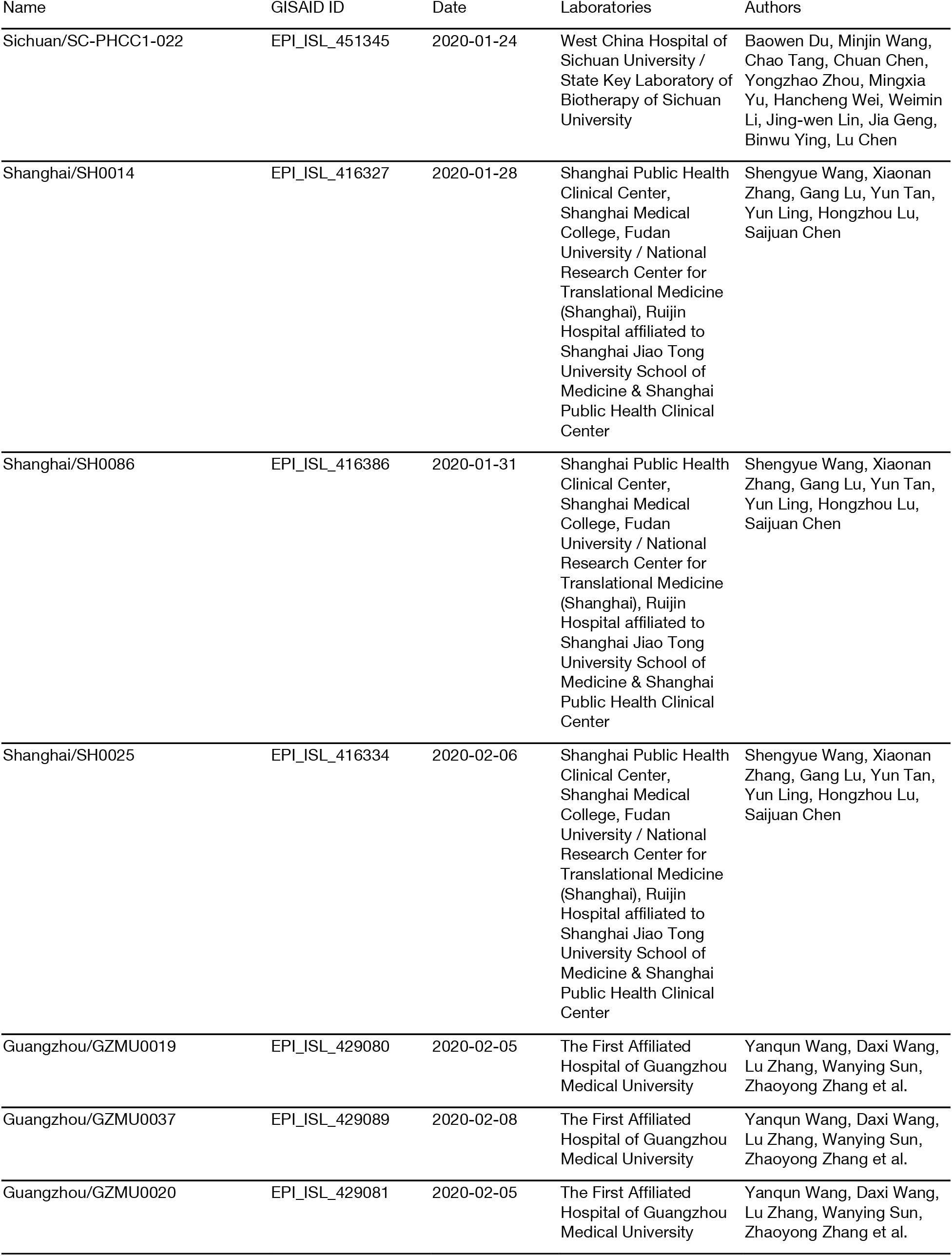
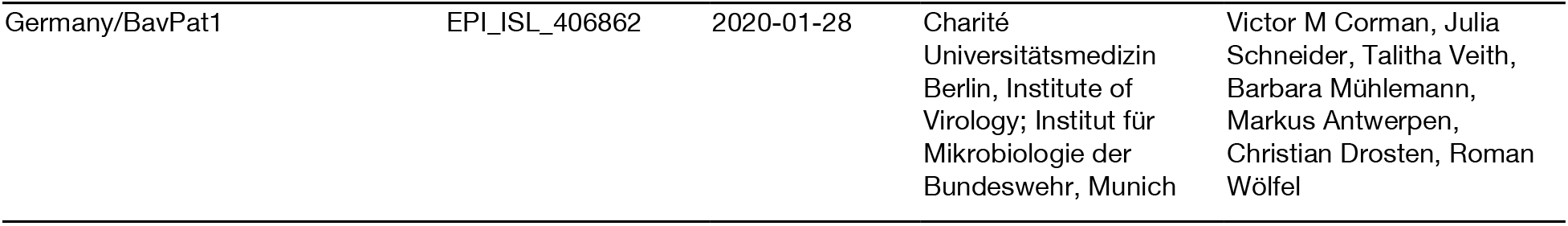
Early D614G viruses in lineage B.

